# Gene-Gene Interactions Between A *LMNA* Variant and Common Polymorphisms Drive Early-Onset Atrial Fibrillation

**DOI:** 10.1101/2025.05.05.25326834

**Authors:** Asia Owais, Hammad Farooq, Hannah Chen, Prisca K Thami, Jaime DeSantiago, Talla Abbas, Arif Pavel, Bradley Merril, James S Ware, Fu Siong Ng, Dawood Darbar

**Author notes:** Address correspondence to: Dawood Darbar, MD Department of Medicine University of Illinois Chicago Chicago, IL 60614.

## Abstract

Atrial fibrillation (AF) is a common arrhythmia with a complex genetic basis, yet the molecular mechanisms linking rare and common variants remain unclear. Using induced pluripotent stem cell-derived atrial cardiomyocytes, we uncover a novel mechanism by which a rare pathogenic *LMNA* variant encoding Lamin A/C disrupts chromatin accessibility and gene regulation at AF-associated loci. Specifically, reduced accessibility at an *SCN5A* enhancer harboring an AF-associated variant leads to reduced sodium current, conduction abnormalities, and re-entrant AF. These electrophysiological defects are rescued by CRISPR-mediated activation of the *SCN5A* promoter and enhancer, providing the first molecular evidence of epistatic gene-gene interactions driving arrhythmia risk and mechanistically linking atrial myopathy and AF. At the population level, we demonstrate that carriers of *LMNA* protein-altering variants with a high polygenic risk score are at a two-fold increased risk of early-onset AF, highlighting the need to integrate rare and common variants for more accurate AF risk assessment.

## Introduction

Atrial fibrillation (AF) is the most common cardiac arrhythmia worldwide and has a strong genetic basis^1^. Both rare and common genetic variants contribute to AF susceptibility^2^. Genome-wide association studies (GWAS) have identified numerous common variants associated with AF, whereas whole genome and whole exome sequencing studies have uncovered individually rare, high-impact variants^3,4^. The development of polygenic risk scores (PRS) has improved AF risk prediction, with emerging evidence suggesting that common variants can modulate the penetrance of rare monogenic variants^4–6^. However, the molecular mechanisms underlying these gene-gene interactions and their role in creating an arrhythmogenic substrate remain poorly understood. A deeper understanding of these epistatic interactions could open new avenues for precision medicine and improve risk stratification.

Variants in *LMNA*, which encodes the nuclear envelope protein Lamin A/C, are implicated in a spectrum of disorders, including isolated cardiac disease^7^. Clinically, *LMNA* mutations are associated with conduction system disease, AF, ventricular arrhythmias, and dilated cardiomyopathy (DCM)^7^. AF is a particularly common and early manifestation in *LMNA*-associated cardiomyopathy^3,8,9^. Lamin A/C, as type V intermediate filaments, play critical roles in cytoskeletal and nuclear structural integrity, chromatin organization, and gene regulation. *LMNA* variants can disrupt gene expression through epigenetic mechanisms, including altered chromatin accessibility and genome organization^4,10–12^.

We hypothesized that a rare *LMNA* variant may alter chromatin accessibility and gene expression at AF-risk loci, unmasking the effects of common polymorphisms in cis-regulatory regions and thereby influencing AF susceptibility in carriers of protein-altering variants (PAVs) in *LMNA*. To test this, we conducted chromatin accessibility and gene expression profiling in induced pluripotent stem cell-derived atrial cardiomyocytes (iPSC-aCMs) generated from a proband carrying the *LMNA*-S143P variant. This analysis identified significantly altered chromatin accessibility at eight AF-associated loci, including a single nucleotide polymorphism (SNP) within a known *SCN5A* enhancer^13^. SCN5A expression and sodium channel current (*I*_Na_) were consistently reduced, impairing atrial conduction and promoting re-entry, thereby predisposing to early-onset AF (defined as AF occurring prior to 66 years)^9,14^. Gene expression changes at additional AF-associated loci identified via Promoter-Capture Hi-C (PC-Hi-C) further reinforced the role of epistatic interactions in AF susceptibility^15^.

To investigate the functional relevance of these chromatin changes, we employed CRISPR-based approaches to correct the *LMNA*-S143P variant, activate the *SCN5A* promoter, and enhance *SCN10A* regulatory activity. These interventions successfully restored SCN5A expression and *I*_Na_, confirming that chromatin remodeling at this AF-risk locus was a key driver of the observed electrophysiological (EP) defects. Our findings support a model in which epistatic interactions between a rare *LMNA* variant and common polymorphisms establish a pro-arrhythmic molecular substrate, contributing to AF pathogenesis.

Prior studies suggest that common variants modulate the penetrance of rare monogenic variants^4,6^. To determine the combined contribution of common polymorphisms and rare *LMNA* variants to AF risk, we applied a PRS for AF to carriers of rare *LMNA* PAVs in the UK Biobank and *All of Us* (AoU) cohorts. Individuals with a high PRS exhibited a two-fold increased risk of early-onset AF compared to those with a lower PRS emphasizing the need to integrate rare and common genetic variations for improved AF risk stratification.

This study unveils a novel molecular mechanism through which epistatic gene-gene interactions drive early-onset AF in *LMNA* variant carriers. These findings highlight the need for comprehensive genetic assessment, incorporating both rare and common variants, not only for risk prediction but also for early intervention to prevent atrial myopathy and its complications, including stroke.

## Results

### Cardiac Phenotype of LMNA-S143P Kindred

The proband (III-3) developed early-onset AF in her 30s. She initially underwent direct current cardioversion to maintain sinus rhythm, but the persistence of symptomatic AF necessitated pulmonary vein ablation a year later. Notably, diffuse low-amplitude atrial electrograms were recorded during the ablation. The patient, however, continued to experience symptomatic persistent AF with bradycardia and underwent implantation of a biventricular pacemaker (**Table 1**). A transthoracic echocardiogram at the time of the ablation showed evidence of mild atrial myopathy but preserved left ventricular function with an ejection fraction of 56%. Cardiac magnetic resonance imaging also showed an enlarged left atrium but no evidence of DCM. A detailed family history revealed that multiple family members were affected by cardiovascular disease (**Fig. 1a**). Most family members started to experience symptoms in their mid-30s. The maternal grandmother (I-2) and several of her siblings died due to congestive heart failure (CHF). Three family members (II-2,II-6 and III-4) developed early-onset AF, while one family member (II-4) underwent heart transplantation due to CHF. The proband’s maternal cousin (III-7) died of sudden death in the 30’s. Several family members have also undergone pacemaker implantation. Genetic testing of the proband confirmed the presence of a heterozygous missense pathogenic variant in the second exon of *LMNA*. A base substitution from Thymine to Cytosine results in an amino acid change from Serine to Proline (NM_170707.4(LMNA):c.427T>C [p.Ser143Pro]) (**Fig. 1b**). The variant is classified as “pathogenic” for DCM by the American College of Medical Genetics and Genomics guidelines^16,17^. It is also the most prevalent variant in *LMNA* associated with DCM in Finland^18,19^. Genetic testing of the affected mother (II-2) confirmed that she also harbored the *LMNA*-S143P variant, while cascade screening determined that the the proband’s sibling (III-2) was not a carrier.

**Fig. 1:**
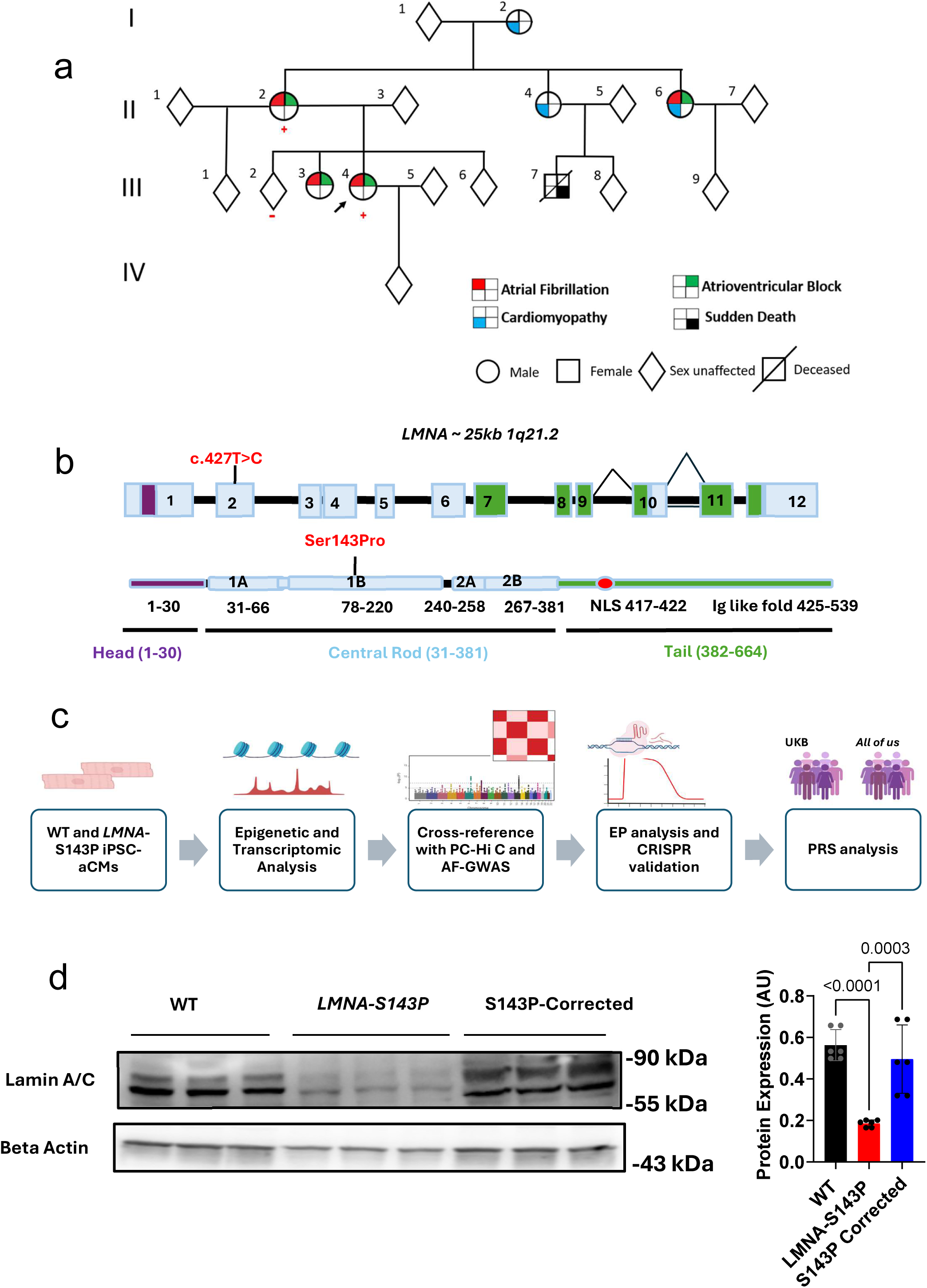
Pedigree of a family harboring the pathogenic *LMNA*-S143P variant. **a.** Schematic representation of the family pedigree, highlighting affected members with a history of atrial fibrillation (AF), atrioventricular (AV) block, cardiomyopathy, and sudden death. The arrow indicates the proband. Circles denote females; squares denote males. The plus (+) and minus (-) signs indicate the presence and absence of the *LMNA*-S143P variant, respectively. **b.** Schematic illustrating the nucleotide and amino acid change (NM_170707.4(LMNA):c.427T>C [p.Ser143Pro]) in exon 2 and the central rod domain. The arrowhead marks an alternative splice site location for generating Lamin A and C transcripts. **c.** Overview of the study design. **d.** Protein expression analysis of Lamin A/C in wild-type (WT), LMNA-S143P, and S143P-corrected iPSC-aCMs.

**Table 1:** Cardiac Phenotype of *LMNA*-S143P Kindred.

|  | AF | Conduction Disease | DCM | SCD | PPM/ICD | Additional Phenotyping |
| --- | --- | --- | --- | --- | --- | --- |
| I-2 |  |  | Yes |  |  | Died in '40s<br>5 out of 7 siblings died in the '40s due to CHF |
| II-2 + | Yes | Yes |  |  | PPM |  |
| II-4 |  | Yes | Yes |  | PPM | Heart transplant |
| II-5 | Yes | Yes | Yes |  | CRT-D |  |
| III-2 - |  |  |  |  |  |  |
| III-3 | Yes | Yes |  |  |  | Bradycardia,<br>1°AVB |
| III-4 + | Yes | Yes |  |  | BiV-PPM | Bradycardia,<br>1°AVB |
| III-7 |  |  |  | Yes |  |  |
| III-8 |  |  |  |  |  | SVT, NSVT |
| III-9 |  |  |  |  |  | POTS |
| III-5 |  |  |  | Yes |  | Died in 30s' |
| III-6 |  |  |  |  |  | SVT, NSVT |
| III-9 |  |  |  |  |  | POTS |

Abbreviations: congestive heart failure (CHF), biventricular pacemaker (BiV-PPM), atrioventricular block (AVB), supraventricular tachycardia (SVT), non-sustained ventricular tachycardia (NSVT), postural orthostatic tachycardia syndrome (POTS) "+" genotype positive, "-" genotype negative

To identify the molecular mechanisms by which epistatic rare-common variant interactions create a substrate for AF, we generated iPSCs from the proband, healthy control, and CRISPR corrected the mutation to generate an isogenic control labeled as *LMNA*-S143P, wild-type (WT), and *LMNA*-S143P-Corrected respectively. The iPSCs were differentiated into atrial cardiomyocytes and matured for four weeks before analysis. Protein expression analysis revealed reduced Lamin A/C levels in *LMNA*-S143P iPSC-aCMs compared to controls, suggesting haploinsufficiency caused by the mutant allele (**Fig. 1d**, Supplementary Figs. 1-2).

### The LMNA-S143P variant causes extensive transcriptional changes and alters regulatory interactions in AF susceptibility genes

Lamins anchor chromatin to the nuclear periphery in transcriptionally inactive lamina-associated domains (LADs), while also interacting with transcriptionally active chromatin within the nucleoplasm^20^. Variants in *LMNA* can disrupt chromatin organization and influence gene expression through epigenetic mechanisms^21^. To investigate the alterations in gene expression due to altered chromatin contacts and accessibility, we performed gene expression and chromatin accessibility profiling using RNA-sequencing (RNA-seq) and Assay for Transposase-Accessible Chromatin (ATAC)-seq of *LMNA*-S143P iPSC-aCMs and WT controls and cross-referenced with published Promoter Capture Hi-C (PC-Hi-C) data from iPSC-derived cardiomyocytes (**Fig. 1c**)^15^.

Differential gene expression (DEG) analysis revealed extensive transcriptional changes, with 4,341 upregulated and 4,015 downregulated genes (**Fig. 2a**). Notably, genes critical for cardiac conduction were significantly altered, with marked downregulation of *SCN5A*, a key regulator of conduction strongly linked to early-onset AF^22,23^ (**Fig. 2b**). Structural genes associated with early onset-AF such *as MYL4, TTN, MYBPC3, DSP* and *PKP2* were also significantly downregulated along with other cytoskeletal genes (**Fig. 2c-d**)^24^ Gene enrichment identified altered cardiovascular signaling pathways, including upregulation of DCM, β-adrenergic signaling and downregulation of the cardiac hypertrophy and differentiation pathways (**Fig. 2e**, Supplementary Figs. 4a-b). The DNA damage repair pathway was also upregulated, alluding to the impact on the nucleus and chromatin due to impaired mechanical stress resilience of the Lamin A/C deficient nuclear lamina **(Fig.1d** Supplementary Fig. 5 and 8).

**Fig. 2:**
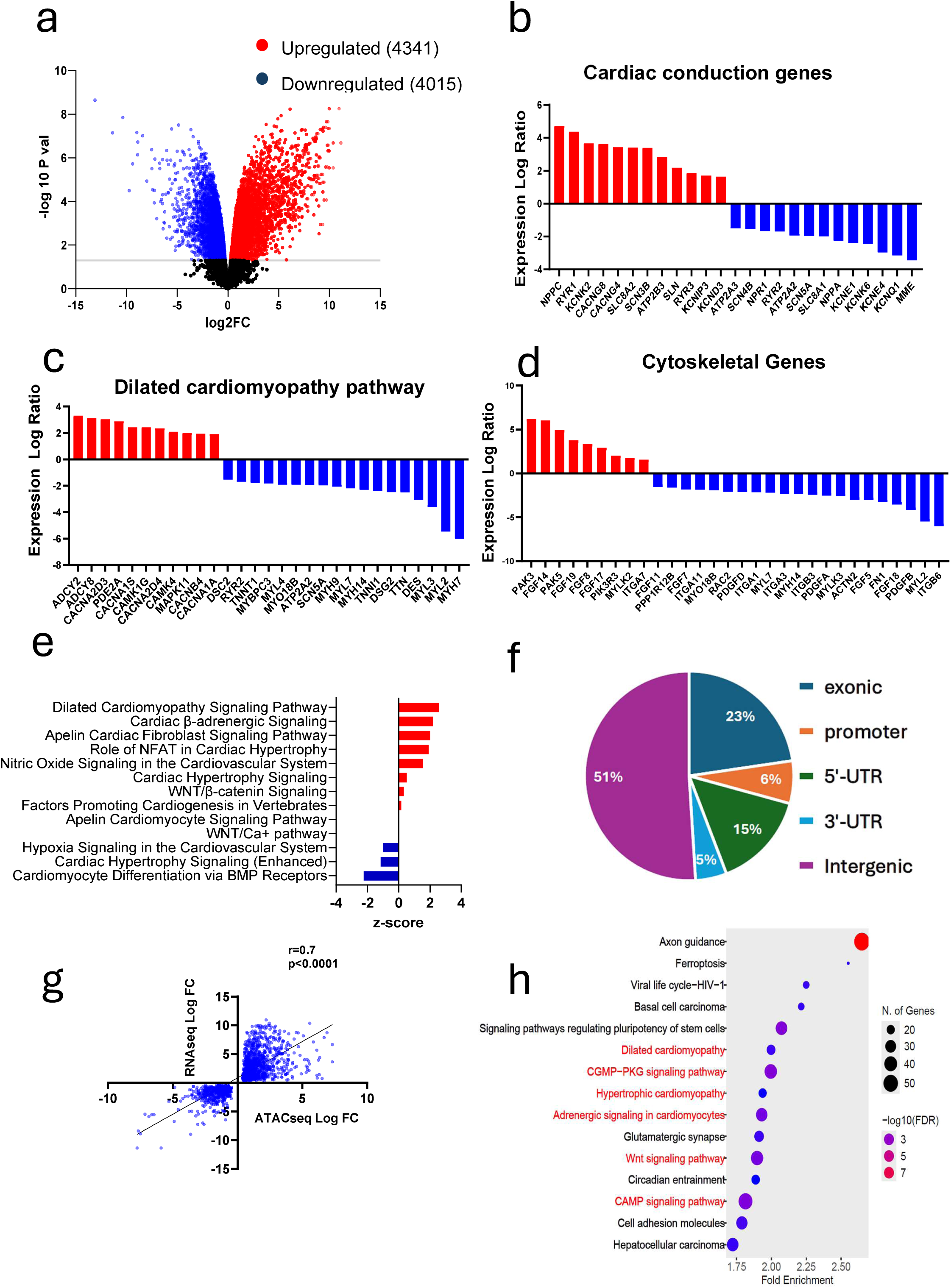
Transcriptomic and chromatin accessibility changes in *LMNA-S143P* iPSC-aCMs. **a.** Volcano plot showing differentially expressed genes (DEGs) (*FDR* < 0.05, *log₂FC* > 0). **b-d.** Differential Gene expression changes in genes associated with dilated cardiomyopathy (DCM), cardiac conduction, and cytoskeletal function. **e.** Altered cardiovascular signaling pathways in *LMNA-S143P* iPSC-aCMs as identified through Ingenuity Pathway Analysis (IPA). **f.** Distribution of differential ATAC-seq peaks, illustrating that the majority are located in non-coding genomic regions. **g.** Positive correlation between gene expression and chromatin accessibility at promoters of differentially expressed genes. **h.** KEGG pathway analysis of differential ATAC peaks at promoter regions.

ATAC-seq-based chromatin accessibility profiling identified 44,981 differential chromatin accessibility peaks between the two groups, with 25,151 upregulated and 19,830 downregulated in *LMNA*-S143P iPSC-aCMs compared to WT. Notably, most of these differential peaks were in non-coding regions (**Fig. 2f**), suggesting that the widespread changes driven by the *LMNA*-S143P variant may result from its impact on cis-regulatory regions, such as enhancers or silencers. Chromatin accessibility changes at transcription start sites (TSS) of the majority (77%) of DEGs correlated positively with gene expression changes (r = 0.75; p<0.0001; **Fig. 2g**). Pathway analysis of genes with altered promoter accessibility revealed significant enrichment in pathways related to cardiac development and repair, including, DCM, calcium signaling, and the Wnt signaling (**Fig. 2h**).

To further investigate changes in chromatin contacts in regulatory regions, we identified enhancers using published H3K27ac chromatin immunoprecipitation (ChIP)-seq data from human atrial tissue (ENCSR074ECR) and mapped their target genes by cross-referencing published PC-Hi C data^15^. Our analysis revealed significant alterations in chromatin contacts between promoters and enhancers in *LMNA*-S143P iPSC-aCMs. We found that there was a strong correlation of enhancer accessibility with differentially expressed target genes (r=0.6856, r^2^=0.47, p<0.0001) (**Fig.3a**). Pathway analysis of the target genes of enhancer regions with altered accessibility showed enrichment for GO terms related to heart development and cytoskeletal reorganization (**Fig. 3b**). We found reduced accessibility at enhancers of genes encoding transcription factors, *TBX5, GATA4, GATA6*, and the atrial-specific gene, *NPPA* encoding atrial natriuretic peptide, a key signaling molecule secreted in response to atrial stretch (**Fig. 3c-g**). The concurrent reduction in the expression levels of these genes suggested altered promoter enhancer contacts. *GATA4, GATA6*, and *TBX5* encode transcription factors that work co-operatively to drive a gene expression program for cardiac development and have been implicated in conduction disorders and familial AF^3,25–29^. *TBX5* maintains an atrial enhancer network regulating atrial-specific genes^30^. We found downregulation of several known TBX5 transcriptional targets encoding ion channels, calcium signaling genes, and structural proteins (**Fig. 3h**)_30-32_.

**Fig. 3:**
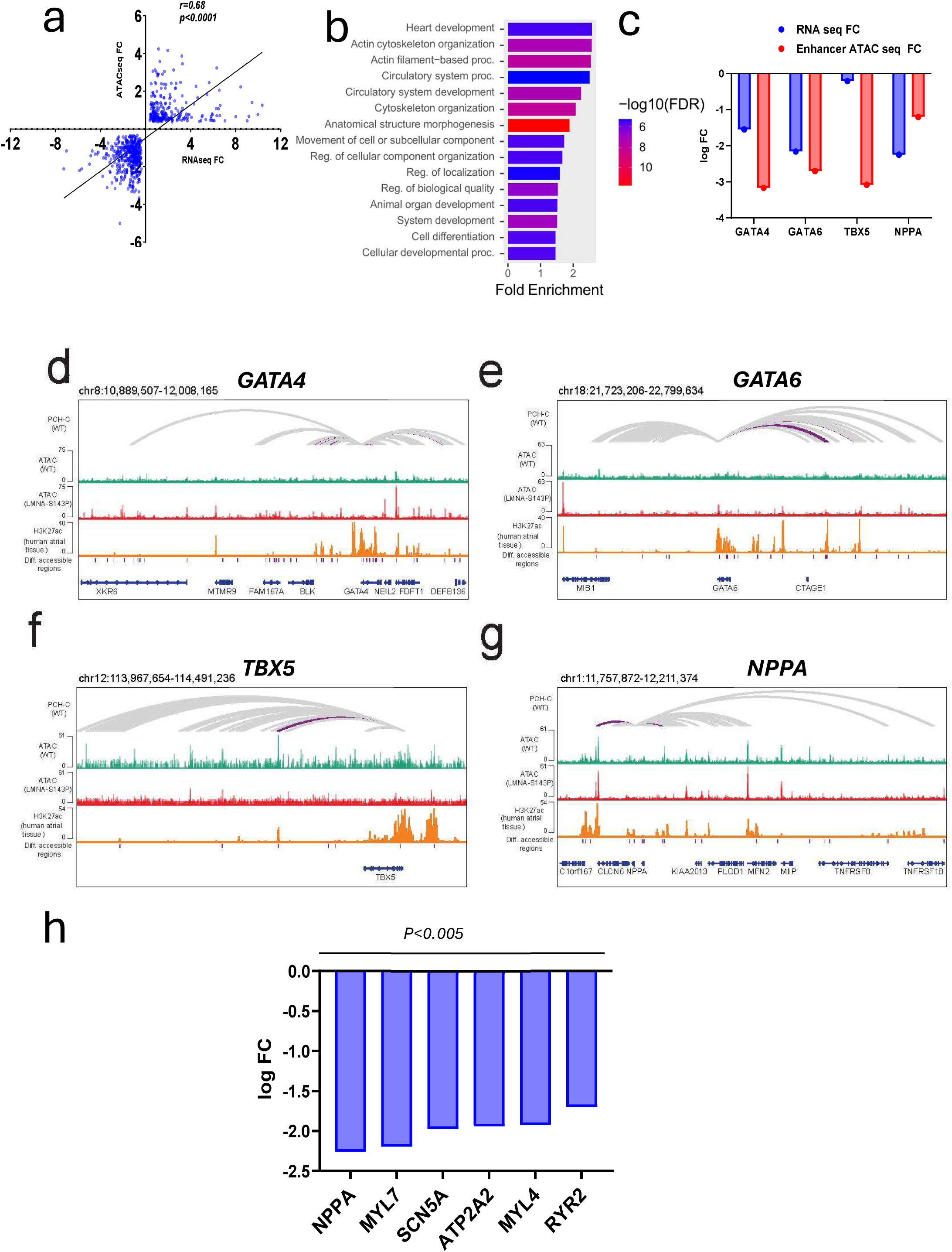
Altered promoter-enhancer interactions in *LMNA-S143P* iPSC-aCMs at AF-associated genes. **a.** Positive correlation between enhancer accessibility and differential expression of target genes. **b.** Gene enrichment analysis of target genes associated with differentially accessible enhancers. **c.** *ATAC*-seq and *RNA*-seq fold changes of target genes linked to differentially accessible enhancers. (*FDR* < 0.05, *log2 FC* > 0 three replicates of each group) **d-g.** Promoter interactions for *GATA4*, *GATA6*, *TBX5*, and *NPPA*. Dark purple arcs indicate enhancer interactions with significant changes in accessibility. **h.** Significant downregulation of *TBX5* transcriptional targets from RNA sequencing.

Overall, these results suggest that the gene expression changes in *LMNA*-S143P iPSC-aCMs are partly driven by altered chromatin contacts in regulatory regions of genes involved in atrial arrhythmogenesis, which may increase susceptibility to AF.

### Chromatin accessibility at AF susceptibility loci is altered by the LMNA-S143P variant in iPSC-aCMs

To further understand the genetic mechanisms that predispose to early-onset AF, we hypothesized that altered accessibility at AF risk loci may modulate the AF phenotype in carriers of rare PAVs in *LMNA* through epistatic interactions. To test this hypothesis, we cross-referenced differential ATAC peaks with AF-associated risk loci identified by three recent AF-GWAS^25,33,34^.

By overlapping GWAS AF risk loci with regions of significantly altered chromatin accessibility, we identified eight AF-associated loci (**Supplementary Table 1**). Notably, there was a significantly reduced accessibility at a region harboring the rs6801957 single nucleotide polymorphism (SNP) on chromosome 3 in the *SCN10A* gene, with *SCN5A* as the neighboring gene (**Fig. 4a, Supplementary Table 1**). *SCN10A* encodes the voltage-gated sodium channel Nav1.8 which is minimally expressed in the heart and is primarily expressed in neurons. *SCN5A-SCN10A* locus is a variant-sensitive region encompassing several variants associated with electrocardiographic traits^13,35^. The rs6801957 SNP is in an intronic region of *SCN10A*, a known enhancer region for *SCN5A*^13^. This suggests that alteration in chromatin accessibility at this locus may alter the promoter enhancer interaction leading to reduced expression of *SCN5A*. Expression analysis by qualitative polymerase chain reaction (qPCR) and Western blot showed reduced *SCN5A* mRNA and protein levels in *LMNA*-S143P iPSC-aCMs (**Fig. 4e, f**, Supplementary Fig. 5). To confirm that SCN5A downregulation resulted from reduced transcription rather than proteolytic degradation, we analyzed the gene expression changes in autophagy and endoplasmic reticulum stress pathways involved in proteolysis. The absence of upregulated genes in these pathways supported that Nav1.5 downregulation occurred at the transcript level (Supplementary Fig. 6).

**Fig. 4:**
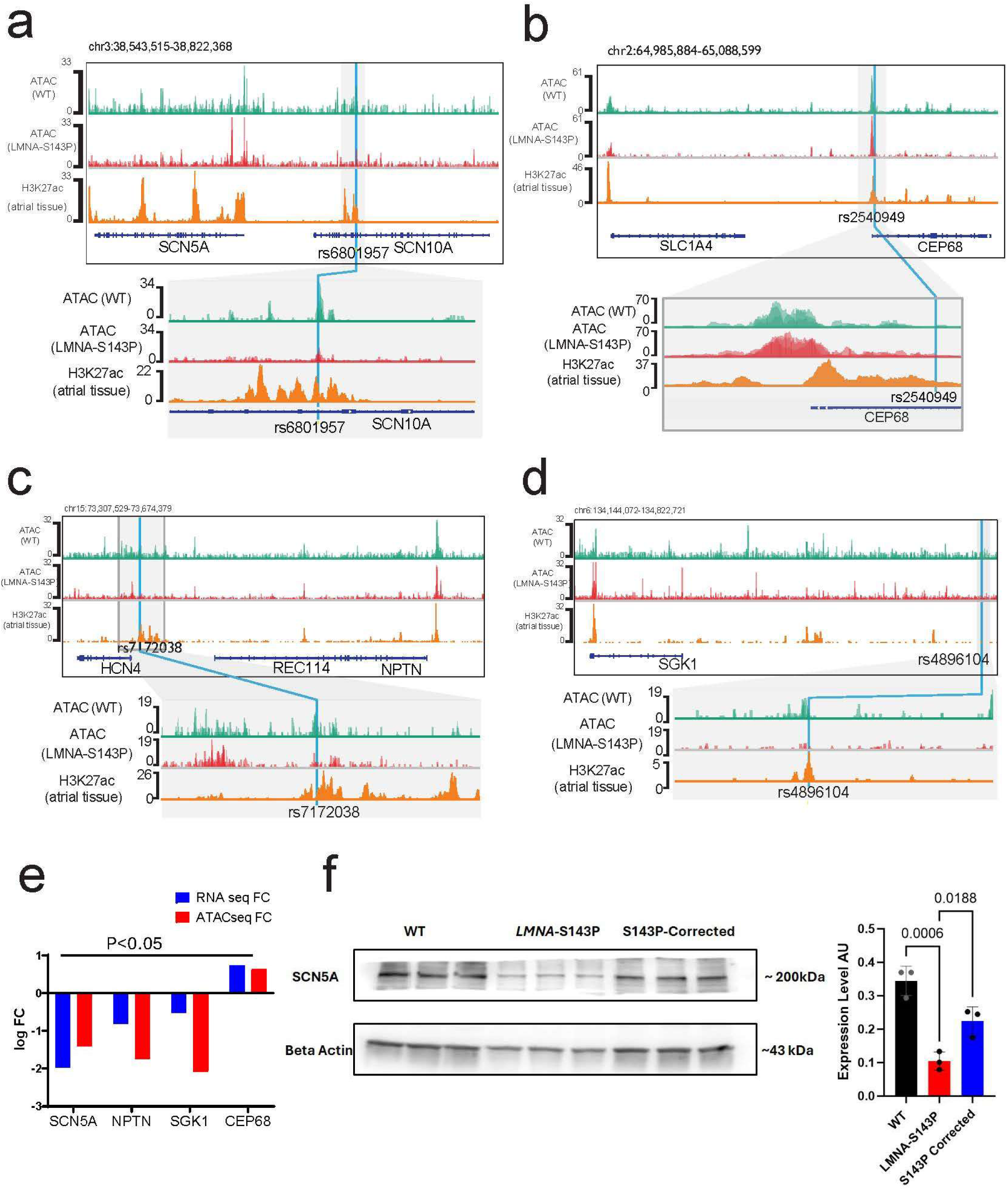
Altered chromatin accessibility in AF risk loci in *LMNA*-S143P iPSC-aCMs. **a-d.** Genome Viewer ATAC and H3K27ac tracks showing altered chromatin accessibility in *LMNA*-S143P iPSC-aCMs at AF-associated loci identified by GWAS in non-coding genomic regions, including enhancer regions harboring SNPs rs6801957, rs2540949, rs7172038, and rs4896104. **e.** Differentially expressed target genes of AF-associated loci overlapping with differential ATAC peaks in regulatory regions. **f.** Validation of *SCN5A* downregulation and rescue by CRISPR correction of the mutation (n=3 representative of three independent differentiations. Ordinary one-way ANOVA with multiple comparisons p<0.05). Significant peaks are indicated at Q < 0.05, log2FC > 0.

Since all the AF-associated SNPs overlapping with differentially accessible regions also overlap with H3K27ac peaks^15^, a marker of active enhancers, we leveraged published PC-Hi-C data from iPSC-derived cardiomyocytes to identify the target genes of these enhancer regions^15^. This analysis revealed altered expression of several target genes implicated in AF pathogenesis (**Fig. 4b-e**). For instance, PC-Hi-C identified *CEP68* as the target gene of the region harboring the rs2540949 SNP. CEP68 was upregulated and is known to play a role in sinus node dysfunction (**Fig. 4b**)^36^. Similarly, Neuroplastin (*NPTN*), identified as the target gene of the region containing rs7172038, is expressed in atrial tissue and involved in atrial differentiation and regulation of heart rate. *NPTN* was significantly downregulated in *LMNA*-S143P iPSC-aCMs (**Fig. 4c**)^37^. Additionally, *SGK1*, encoding Serum and Glucocorticoid-regulated Kinase-1, was significantly downregulated. *SGK1* was identified as a target of an intergenic region harboring the rs4896104 SNP (**Fig. 4d**). This serine-threonine kinase regulates SCN5A activity and has been implicated as a therapeutic target for cardiac arrhythmias^38,39^. These findings suggest epistatic interactions in multiple genomic regions regulating distinct and overlapping pathways may culminate in AF.

### EP Alterations in LMNA-S143P iPSC-aCMs

To assess the electrophysiological (EP) profile of *LMNA*-S143P iPSC-aCMs^40,41,42^, we measured evoked action potentials (AP) in isolated mature iPSC-aCMs. *LMNA*-S143P iPSC-aCMs exhibited altered AP profiles, including reduced AP amplitude and upstroke velocity (dV/dTmax), a key indicator of atrial conduction, while AP duration at 90% repolarization (APD90) remained unchanged (**Fig. 5a-d**). AP amplitude and upstroke velocity are primarily determined by the peak *I*_Na_, conducted via the main cardiac sodium channel Nav1.5, encoded by *SCN5A*. Consistently, voltage-clamp recordings revealed a significant reduction in peak *I*_Na_ density in *LMNA*-S143P iPSC-aCMs compared to WT and *LMNA*-S143P-CRISPR-corrected cells (**Fig. 5e-f**). Stimulated multielectrode array recordings further demonstrated marked conduction heterogeneity across *LMNA*-S143P iPSC-aCM monolayers, a pattern absent in WT and corrected iPSCs (**Fig. 5g**). Importantly, potassium (*I*_K_) and calcium (*I*_Ca_) currents, which regulate the repolarization phase of the AP, remained unchanged, reinforcing that the observed EP changes in *LMNA*-S143P iPSC-aCMs are primarily driven by alterations in *I*_Na_ (Supplementary Fig. 7). In summary, reduced conduction velocity due to *SCN5A* downregulation in *LMNA*-S143P iPSC-aCMs promotes re-entry, establishing an arrhythmogenic substrate for early-onset AF.

**Fig. 5:**
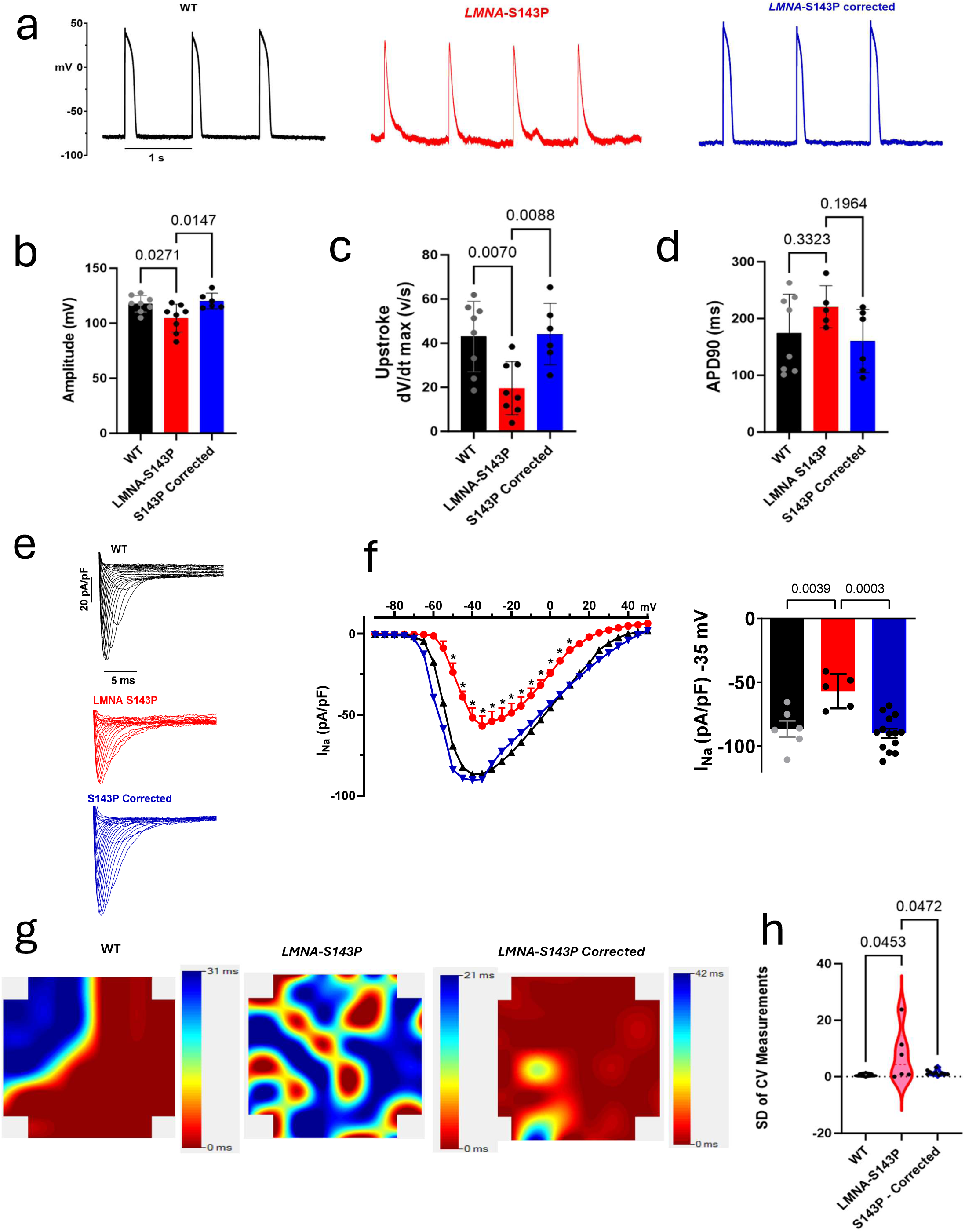
Electrophysiological alterations in *LMNA-S143P* iPSC-aCMs. **a-b.** Representative evoked action potential (AP) traces measured by whole-cell patch clamping show reduced AP amplitude in *LMNA-S143P* iPSC-aCMs, which is rescued by *LMNA-S143P* corrected iPSC-aCMs. **c.** Reduced upstroke velocity/instantaneous rate of voltage change over time (dV/dTmax), an indicator of atrial conduction velocity, in *LMNA-S143P* iPSC-aCMs. **d.** No change in action potential duration at 90% repolarization (*APD90*). **e.** Representative sodium current (*I_Na_*) traces show reduced sodium current density in *LMNA-S143P* iPSC-aCMs. **f.** The current-voltage (*I/V*) curve shows reduced *INa* in *LMNA-S143P* iPSC-aCMs, quantified in the bar graph. (*a-d*, n = 5-7 cells per group, from three independent differentiations). (*Ordinary one-way ANOVA with multiple comparisons p<0.05*). **g.** Activation maps of propagation of electrical activity measured by a multielectrode array (MEA) show marked conduction heterogeneity in *LMNA-S143P* iPSC-aCMs (n = 6 stimulated conduction velocity [CV] measurements for each group). **h.** Increased standard deviation (SD) of conduction velocity measurements in *LMNA-S143P* iPSC-aCMs. (Ordinary one-way ANOVA with multiple comparisons p<0.05).

### Altered Calcium Handling and contractility in LMNA-S143P iPSC-aCMs

Calcium transient measurements revealed that *LMNA*-S143P iPSC-aCMs exhibited reduced transient amplitude and duration and irregular transient intervals (**Fig. 6a-d**). Contractility assessments further demonstrated a decrease in contractile amplitude in *LMNA*-S143P iPSC-aCMs compared to controls (**Fig. 6f**). Transmission electron microscopy identified key myopathic features, including nuclear deformities, myofibrillar disarray and disrupted intercalated discs, consistent with histopathological findings in Lamin A/C cardiomyopathy ^43^(**Fig. 6g-i**). Collectively, these disruptions in calcium handling and contractility support the electromechanical dysfunction underlying atrial myopathy and AF^44^.

**Fig. 6:**
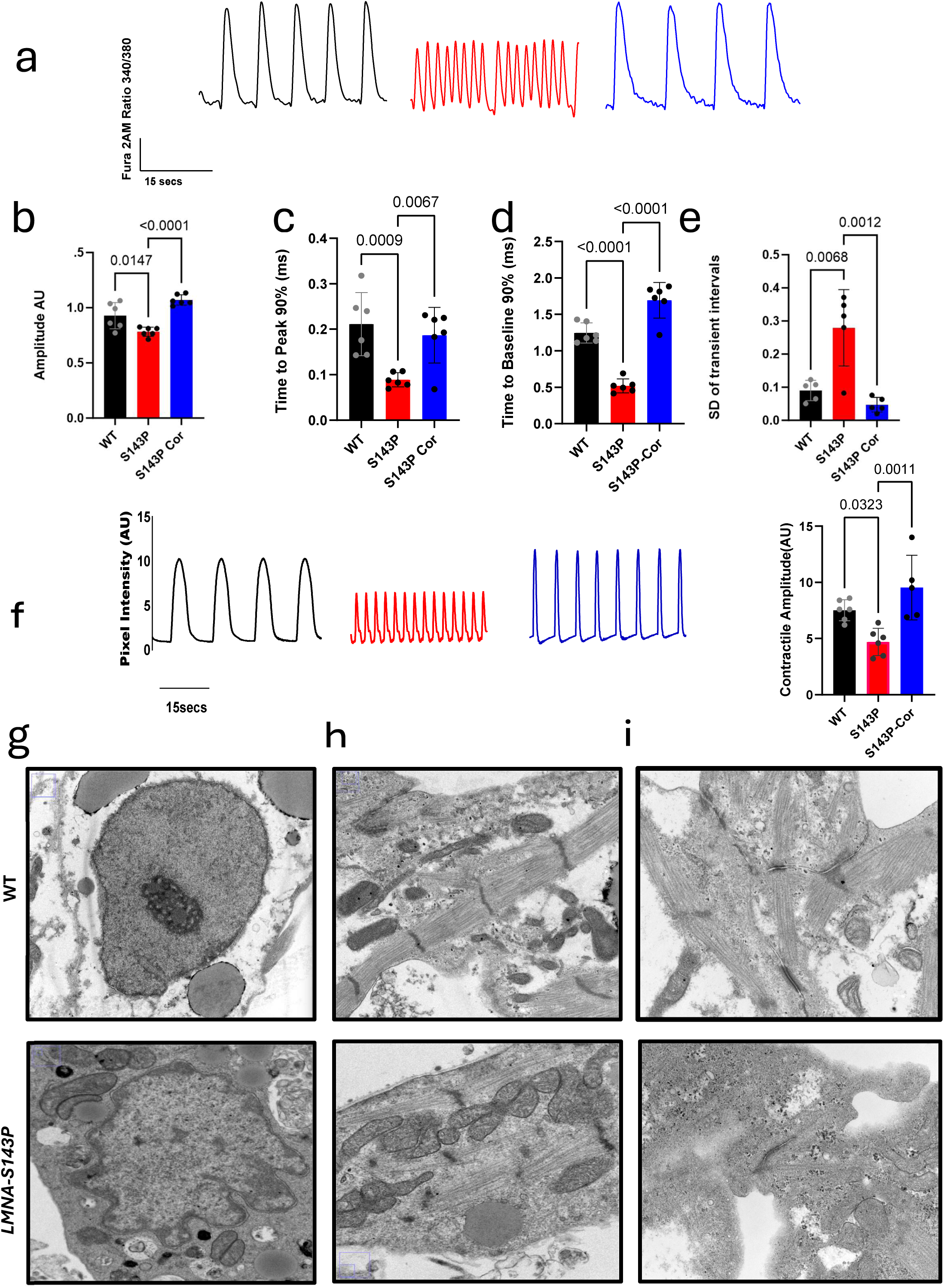
Calcium transient aberrations and decreased contractility in *LMNA-S143P* iPSC-aCMs. **a-e.** Representative traces of calcium transients showing reduced amplitude, shorter time to peak, delayed time to relaxation, and irregularity of transient intervals in *LMNA-S143P* iPSC-aCMs, which are rescued by CRISPR correction of the mutation. **f.** Reduced contractile amplitude in *LMNA-S143P* iPSC-aCMs. **g-i.** Transmission electron microscopy images showing features of atrial myopathy in *LMNA-S143P* iPSC-aCMs. **g.** Pleiomorphic nuclei, wider intermembrane space, and more heterochromatic foci (Scale = 1 μm, 12,000x magnification). **h.** Disorganized sarcomeres and thinner myofibrils (Scale = 1 μm, 15,000x magnification). **i.** Disrupted intercalated discs (Scale = 500 nm, 30,000x magnification). **b-f** n=4-6 recordings representative of three independent differentiations, Ordinary one-way ANOVA with multiple comparisons, p<0.05.

### CRISPR activation of SCN5A restores sodium current density in LMNA-S143P iPSC-aCMs

Electrophysiological analysis suggested that reduced *I*_Na_ in *LMNA*-S143P iPSC-aCMs may result from a closed chromatin conformation at the *SCN5A-SCN10A* locus. We employed a nuclease-deficient Cas9 (dCas9) fused to transcriptional activators Vp64, p65, and Rta to upregulate SCN5A transcription to test this hypothesis. For the *SCN10A* enhancer region, we utilized a specific transcriptional activator, p300, which possesses histone acetyltransferase activity, facilitating chromatin remodeling and enhancer activation^45^. Transfection of both CRISPR activation complexes successfully restored *SCN5A* expression and *I*_Na_ in *LMNA*-S143P iPSC-aCMs, supporting our hypothesis that reduced chromatin accessibility at this locus contributes to *I*_Na_ deficiency (**Fig. 7a-e**). These findings establish a genomic mechanism underlying a pro-fibrillatory, re-entrant substrate driven by reduced *I*_Na_.

**Fig. 7:**
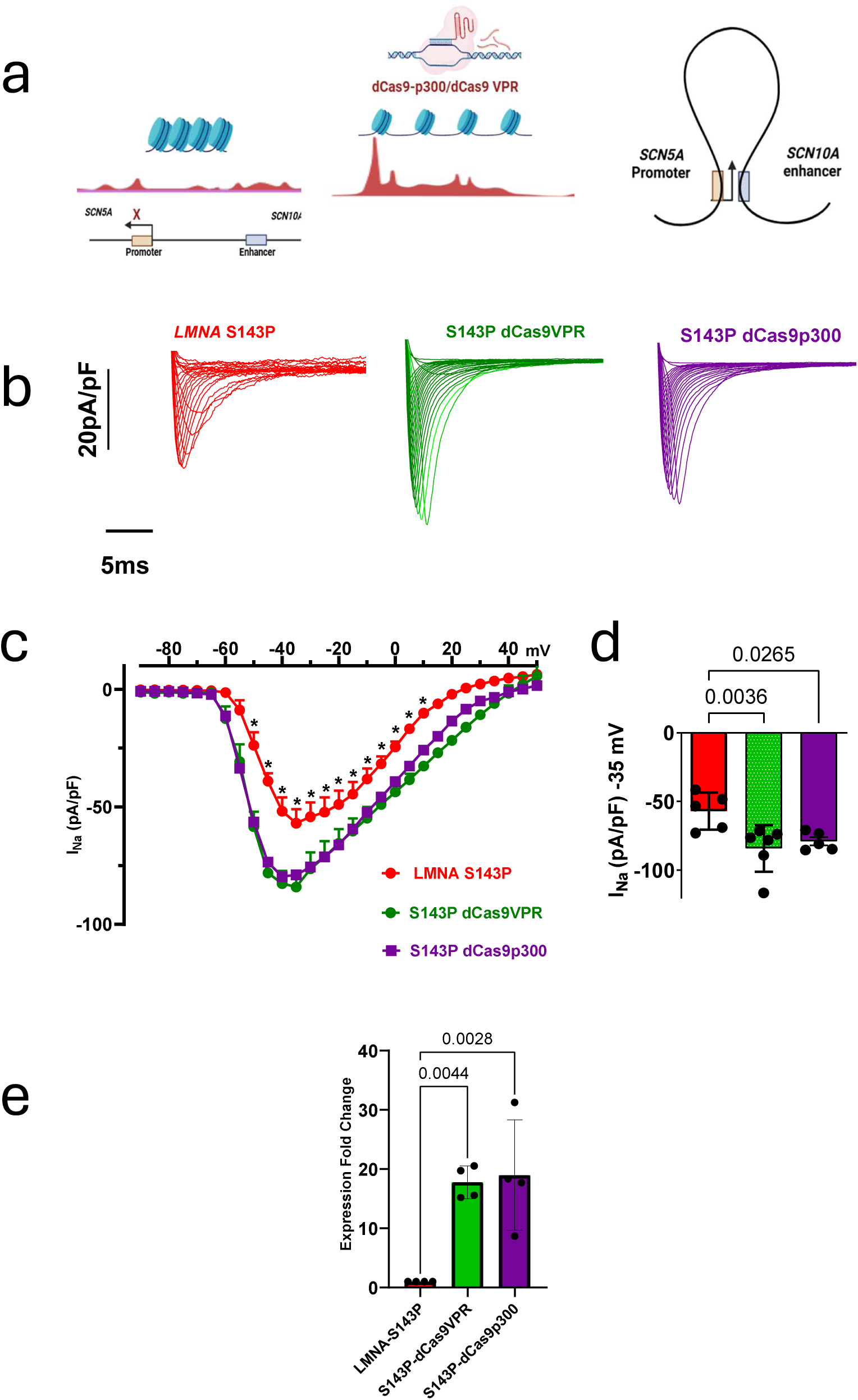
CRISPR activation of SCN5A restores INa in *LMNA-S143P* iPSC-aCMs. **a.** Schematic showing the CRISPR activation design. A closed chromatin conformation at the SCN5A-SCN10A locus inhibits transcription of SCN5A. RNA-guided binding of dCas9-VPR at the SCN5A promoter and dCas9-p300/CBP histone acetyltransferase at the SCN10A enhancer region initiates transcription of SCN5A. **b.** Representative INa traces show restoration of sodium current after CRISPR activation. **c.** Current-voltage (I/V) curve showing reduced INa in *LMNA-S143P* iPSC-aCMs, rescued by transfection of CRISPR activation complexes. **d.** Current-voltage (I/V) curve quantified in a bar graph. (n = 4-6 cells per group; unpaired t-test with Welch’s correction p<0.05). **e.** CRISPR activation restores expression of *SCN5A*; bar graph quantifying changes in expression by qPCR. n=4 replicates representative of three independent differentiations, ordinary one way ANOVA with multiple comparisons, p<0.05)

### Risk of Incident AF in carriers of LMNA protein-altering variants according to PRS

To assess the contribution of common variants to the variable penetrance of early-onset AF in Lamin A/C cardiomyopathy^9^ we applied a PRS for AF to carriers of PAVs, including missense, frameshift, stop-gain, splice, and in-frame insertion variants in *LMNA*, using data from the UK Biobank and *All of Us* (AoU) cohorts. In the UK Biobank, we identified 1,109 carriers of 296 distinct rare *LMNA* PAVs, with an age distribution ranging from 40 to 70 years (median 58 [Q1-Q3: 50-64]), of whom 613 were female. AF prevalence was higher among *LMNA* PAV carriers (n = 89, 8.03%) compared to non-*LMNA* carriers (n = 20,917, 4.99%) (*p* = 5.08 × 10⁻⁶). In non-*LMNA* carriers, a high PRS was significantly associated with an increased risk of AF (hazard ratio [HR] 4.11, *p* < 2 × 10⁻¹⁶) (Supplementary Fig. 9a, Supplementary Table 2). However, the presence of rare *LMNA* PAVs conferred a two-fold increased risk of AF (HR 7.18, *p* < 5.15 × 10⁻⁵), after adjusting for the first 10 genetic principal components, sex, and hypertension (**Fig. 8a, Supplementary Table 3**).

**Fig. 8:**
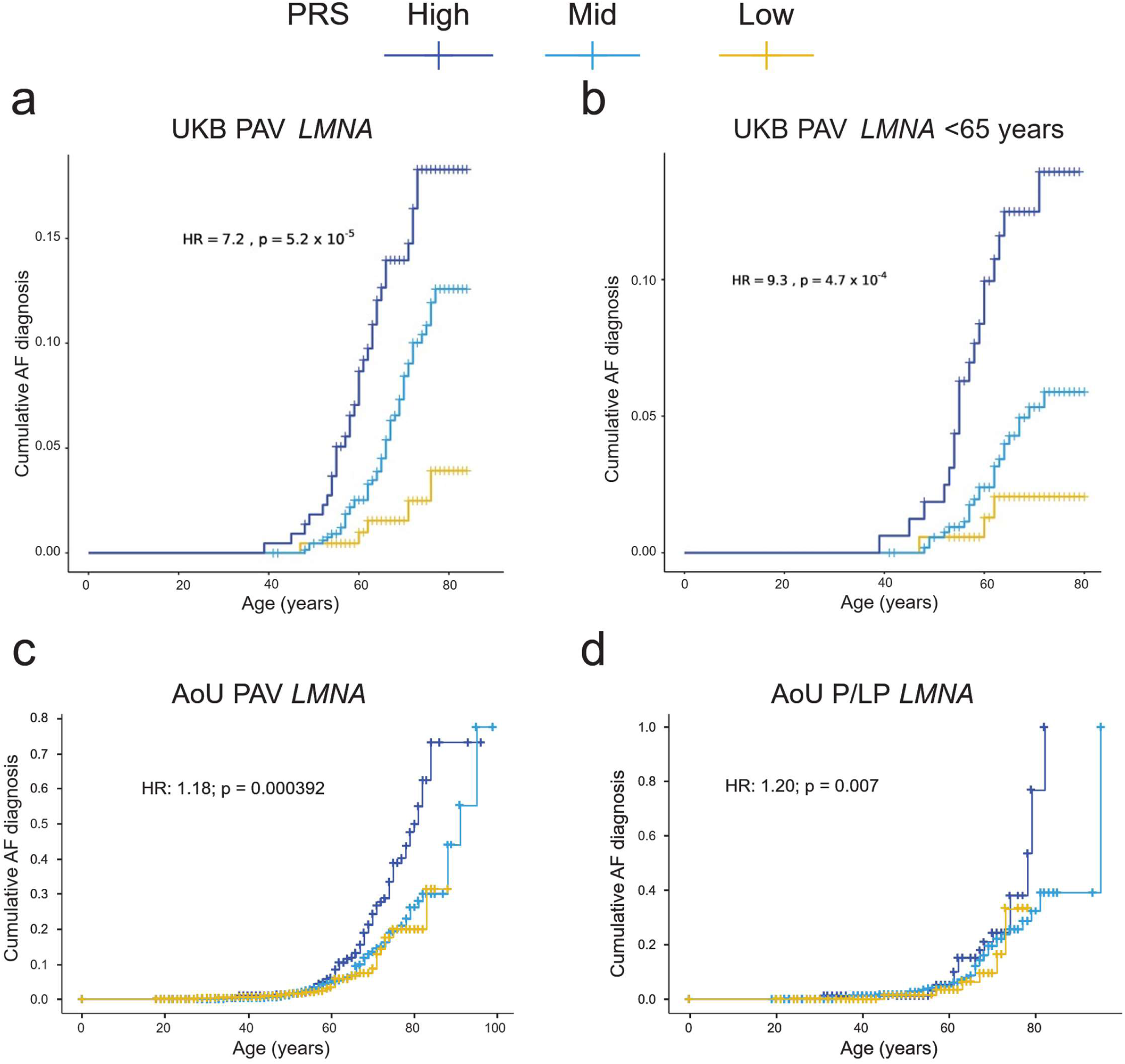
Risk of AF in the UK Biobank (UKB) and All of Us (AoU) cohorts by polygenic risk score (PRS) category. **a** Risk of AF in carriers of protein-altering variants (PAVs) in *LMNA* in the UK Biobank by PRS category (n = 1,109). The risk of AF is doubled in *LMNA* PAV carriers compared to non-LMNA PAV carriers (Supplementary Fig. 9a). **b.** Risk of AF in young (<65 years) participants in the UKB with PAVs in *LMNA* by PRS category is doubled compared to non-*LMNA* PAV carriers (Supplementary Fig. 9b) **c.** Risk of AF in carriers of PAV in *LMNA* by PRS category in the AoU cohort (n = 2,511). A higher PRS is significantly associated with an elevated risk of AF in cases (those with AF) compared to controls (those without AF). **d.** Risk of AF in carriers of Pathogenic/Likely Pathogenetic (P/LP) *LMNA* variants in the AoU cohort by PRS category (n = 510). A significantly higher risk is observed in cases.

We evaluated the association between *LMNA* variants and AF using logistic regression, adjusting for the first 10 genetic principal components, sex, age at recruitment, and hypertension in individuals under 65. *LMNA* variant carriers had significantly higher odds of AF (odds ratio [OR] 1.57, 95% confidence interval [CI] 1.12–2.11, *p* < 4.33 × 10⁻³) compared to non-*LMNA* carriers. The odds increased substantially to 8.17 (CI 2.29–29.2, *p* < 1.23 × 10⁻³) in *LMNA* carriers with a high AF PRS compared to those with a low PRS. In a time-to-event analysis, the cumulative risk of AF was significantly higher in younger *LMNA* carriers than in non-carriers (hazard ratio [HR] 1.56, *p* = 3.01 × 10⁻³) **(Supplementary Table 4)**. High PRS further amplified AF risk, with *LMNA* carriers exhibiting a nearly two-fold greater risk (HR 9.31, *p* < 4.73 × 10⁻⁴) compared to non-*LMNA* carriers (HR 4.4, *p* < 2 × 10⁻¹⁶) in the UK Biobank (**Fig. 8b and Supplementary Fig. 9b &10, Supplementary Tables 5-6**).

We further analyzed 37 carriers of *LMNA* variants classified as pathogenic/likely pathogenic (P/LP) for DCM in ClinVar, of whom six had AF. Four of these six individuals had a high-risk AF PRS (**Supplementary Table 7**). Notably, only one carrier with a P/LP variant had DCM, and this individual also had a high-risk AF PRS. However, due to the limited sample size of P/LP *LMNA* carriers, we could not calculate precise risk estimates for this group.

In the AoU database, we identified 2,511 carriers of *LMNA* PAVs, with an age range spanning 19 to 102 years (median age: 57). AF prevalence was higher among *LMNA* PAV carriers (7.9%) compared to non-*LMNA* carriers (6.18%). Additionally, *LMNA* PAV carriers had significantly increased odds of AF (OR 1.31, 95% CI 1.21–1.51, *p* < 5.38 × 10⁻⁴) relative to non-carriers. Stratification by AF PRS revealed that *LMNA* carriers in the high PRS group had significantly greater odds of AF than those in the low PRS group (OR 3.39, 95% CI 2.10–5.48, *p* < 1.07 × 10^⁻⁷^). Time-to-event analysis using Cox Proportional Hazards modeling, adjusted for the first 10 genetic principal components, sex, and race, further confirmed that a high PRS was associated with an elevated risk of AF (HR 1.18, *p* = 0.000392) (**Fig. 8c**).

To further investigate, we analyzed a subset of 510 *LMNA* PAV carriers with P/LP variants annotated in ClinVar, of whom 54 had AF. Within this P/LP carrier group, individuals with a high PRS had higher odds of AF than those with a low PRS (OR 2.7, 95% CI 1.02–7.42, *p* = 0.06). Consistently, time-to-event analysis confirmed that a high PRS was associated with an elevated risk of AF in this subset as well (HR 1.20, *p* = 0.007) (**Fig. 8d**).

## Discussion

Large-scale exome-wide genetic studies have linked rare *LMNA* variants to early-onset AF^3^; however, the phenotypic expression and severity of AF among carriers remain highly variable. While pathogenic *LMNA* variants associated with DCM exhibit high penetrance, the final clinical phenotype is influenced by the combined effects of multiple variants of a range of effect sizes, contributing to the heritability of complex diseases like AF. Although a PRS quantifies the cumulative impact of common variants, it does not fully capture epistatic interactions, which may be non-additive and context-dependent^46–48^.

By integrating gene expression, chromatin accessibility, and PC-Hi-C genomic interactions, we identified dysregulated regulatory networks in key AF susceptibility genes and regions harboring AF-associated SNPs. These findings reveal a novel molecular mechanism by which rare and common variant epistatic interactions contribute to arrhythmia pathogenesis. Specifically, we observed chromatin accessibility changes at AF-associated loci, including a variant-sensitive *SCN5A* enhancer. These alterations reduced *SCN5A* expression, diminished *I*_Na_, and conduction abnormalities, creating a substrate for re-entrant AF.

To assess the clinical impact of these interactions, we applied an AF PRS in two large population cohorts, the UK Biobank and AoU. Carriers of rare *LMNA* variants with a high PRS had a two-fold increased risk of early-onset AF compared to those with a lower PRS, particularly in individuals under 65. While prior studies suggest that common variants influence the penetrance of rare monogenic variants, our findings provide direct molecular evidence supporting an interaction effect ^4,6^.

Although multiple arrhythmogenic mechanisms are implicated in Lamin A/C cardiomyopathy, such as fibrosis, and gap junction remodeling, we did not find differential changes in regulatory interactions for genes involved in these processes. However, the reduced *I*_Na_ and observed genomic alterations at the *SCN5A-SCN10A* locus strongly suggest that common polymorphisms at this locus increase the genetic predisposition for early-onset AF.

Most AF risk loci identified through GWAS reside in non-coding regions with regulatory roles in gene expression. We observed altered chromatin accessibility at several AF-associated loci and their target genes. While a SNP within the *SCN10A* intronic region had the strongest effect, we also identified associations with *SGK1*, a kinase regulating ion channel activity; *NPTN*, a structural gene involved in atrial differentiation and heart rate regulation; and *CEP68*, which contributes to centrosome cohesion and is linked to sinus node dysfunction^36,37,39^. Although *SCN5A* downregulation emerged as the primary driver of arrhythmic risk, our findings suggest that AF results from broader disruptions involving ion and non-ion channel proteins, including regulatory enzymes and structural components.

To establish a causal link between chromatin remodeling and AF risk, we employed CRISPR activation to restore chromatin accessibility at the variant-sensitive *SCN5A-SCN10A* locus, a region strongly associated with AF and electrocardiographic traits^13,25,49^. This intervention successfully rescued sodium channel expression, demonstrating that interactions between common polymorphisms and sodium channel dysfunction contribute to early-onset AF in *LMNA* variant carriers. Sodium channel dysfunction in individuals with *LMNA* variants has been previously reported in patients exhibiting type I Brugada-like EKG patterns following cardiac arrest, a condition commonly associated with impaired sodium channel activity^50^. Our study provides a mechanistic explanation for this association and supports the inclusion of *LMNA* in genetic testing panels for Brugada syndrome.

Several limitations of this study warrant consideration. While human cardiac tissue represents the ideal model for investigating AF pathophysiology, its limited availability necessitates using iPSC-aCMs as an alternative^51,52^. A recognized limitation of iPSC-derived models is their relative immaturity. To address this, we implemented a comprehensive maturation protocol that combines electrical and biochemical stimuli to mimic fetal and postnatal development^40^. This approach improved the structural and metabolic maturity of iPSC-aCMs, as demonstrated in our previous work modeling an *NPPA* variant, where we identified mitochondrial dysfunction and ion channel remodeling^40^. Compared to animal models, which exhibit species-specific differences in ion channel expression, iPSC-aCMs more accurately recapitulate the diversity of human atrial ion channels. Additionally, iPSC-aCMs offer a unique advantage by allowing the study of pathogenic mechanisms within a patient’s genetic background, providing insights that other experimental models may not fully capture, particularly in assessing the cumulative effects of multiple common polymorphisms. Another limitation of this study is that our exploration of epistatic interactions was based on iPSC-aCMs derived from a single proband. While this approach provides functional evidence for how common variants modulate the variable expressivity of early-onset AF in *LMNA* variant carriers, future studies incorporating additional AF-linked rare variants will be essential to generalize these findings and further elucidate epistatic mechanisms.

The findings of this study have several important implications. While ion channel dysfunction plays a central role in arrhythmia pathogenesis, our results highlight the significance of chromatin remodeling at multiple AF-associated loci. This underscores the complex etiology of AF and the need to expand therapeutic strategies beyond traditional ion channel-targeting drugs. The interaction between a structural gene variant and an AF susceptibility locus enriched with variants associated with electrocardiographic traits establishes a direct link between atrial myopathy and AF. This suggests that screening for atrial myopathy, particularly in family members of affected individuals, may have prognostic value, especially in deciding which patients to select for anticoagulation therapy^53^. Additionally, our successful CRISPR-based activation of the *SCN5A-SCN10A* region demonstrates the potential of epigenetic modifiers as targeted therapeutic interventions for AF, particularly in *LMNA* variant carriers. Finally, our findings highlight the necessity for improved genetic risk prediction models for AF and other complex diseases. Such models should integrate the cumulative effects of common variants with small effect sizes alongside highly penetrant rare variants while accounting for epistatic interactions.

## Methods

### Recruitment of participants

We used the UIC Institutional Review Board–approved protocol to enroll participants after receiving informed written consent.

### Generation of human iPSCs and differentiation to atrial cardiomyocytes iPSC-aCMs

Human iPSCs were derived from reprogrammed peripheral blood mononuclear cells (PBMCs) from the proband and unaffected controls, as previously described^54^. iPSCs were differentiated using the STEMdiff atrial cardiomyocyte differentiation kit. The atrial cardiomyocyte population was purified through glucose starvation and lactate replacement, resulting in contracting monolayers of iPSC-aCMs. Human iPSC-aCMs were then matured using the maturation protocol following dissociation and replating on fibronectin-coated plates and maintained in Cardiomyocyte Maintenance Media supplemented with T3, insulin-like growth factor-1, and dexamethasone as previously described^40,42^. Our protocol typically yields ∼80 to 90% pure iPSC-aCMs and fibroblasts based on immunostaining analysis, as we have previously described^40,42,54^.

### Generation of isogenic control from LMNA-S143P by CRISPR correction

As previously described, the isogenic control (LMNA-S143P Corrected) was generated using the Alt-R CRISPR-Cas9, homology-directed repair (HDR) system. ^40^. Single guide (sg)RNAs and HDR oligos were nucleofected in *LMNA*-S143P hiPSCs. The sgRNAs were designed using the IDT CRISPR design tool based on an algorithm predicting optimal off-target and on-target scores. sgRNA 5’GGAGGCTCTGCTGAACCCCA-3’ was used for genetic correction of Pro143 to Ser, C to T in exon 2. Single-cell clones were isolated, expanded, and verified by next-generation sequencing and Sanger sequencing **(Supplementary Fig. 3b**).

### Electrophysiology

Whole-cell-patch-clamp recordings on single-cell iPSC-aCMs to measure INa, ICa, L, and IK were performed according to previously published protocols using the Axopatch 200B amplifier controlled by pClamp10 software through an Axon Digidata 1440A^40,42,54^. Sodium current measurements were done in a voltage clamp setup; pipettes were filled in mM: 135 CsF, 5 NaCl,5 MgATP, 10 Hepes, 10 EGTA, pH 7.2 with CsOH. Cells were perfused with a solution in mM: 85 CsCl, 10 glucose, 1 MgCl2, 1 CaCl2, 20 Hepes, 50 NaCl, pH 7.4 with CsOH.; 10 uM Nifedipine was added to block calcium channel. Sodium current was elicited by a series of 600 ms test potentials varying from −90 to −50 mV in 5 mV increments from a holding potential of −120 mV.

L-type calcium current was obtained in the whole cell voltage clamp configuration. External recording solution contained (in mM) 136 TEA-Cl, 2 CaCl2, 1.8 MgCl2, 10 HEPES, 5 4-aminopyridine, and 10 glucose (pH 7.4 with TEA-OH). The pipette solution contained (in mM): 125 CsCl, 20-TEA-Cl, 10 EGTA, 10 HEPES, 5 phosphocreatine, 5 Mg2ATP (pH 7.2 with CsOH). IPSC-aCM were held at −80 mV, and 10 mV depolarizing steps from −50 mV to + 50 mV for 300 ms were applied. Recording solutions allowed for the specific measurement of the calcium current without the contaminating Na and K currents.

Potassium current was measured in a whole cell rupture voltage clamp with patch clamp pipettes filled with internal solution in mM: 100 K-aspartate, 20 KCl, 5 Mg ATP, 2 MgCl2, 10 HEPES, 5 EGTA pH 7.2 with KOH; External solution was normal tyrode solution in mM: 140 NaCl, 4 KCl, 1.2 MgCl2, 1.8 CaCl2, 10 glucose, 10 HEPES pH 7.4 with NaOH; 10 uM Nifedipine was added to block calcium channels. IK outward current was measured from a holding potential of −60 mV to test potentials of –30 mV to +50 mV with 20 mV increments for 3 seconds.

### Calcium transient recordings and Contractility Measurements

The Ionoptix system was used for contractility and calcium transient measurements. Contracting monolayers of iPSC-aCMs were incubated in 1uM Fura 2 AM calcium binding dye dissolved in Tyrode’s solution. After 20 min of incubation, the dye solution was removed and replaced with Tyrode’s solution. Calcium transients were analyzed using the ION-OPTIX analysis software. The software performs background detection and generates traces after subtraction of the background. Cytomotion, a label-free detection system, was used for contractility analysis, which provides high-speed data acquisition at 1000Hz. Differences in contractile amplitude measured inotropic differences. Contractility and calcium transient analysis by the ION wizard software is done according to an algorithm optimized for iPSC-derived cardiomyocytes.

### Conduction Velocity Recordings

iPSC-aCMs from the three groups were plated on dishes with multi-electrode arrays. Biphasic stimuli were applied to iPSC-aCMs at an amplitude of 1000mV for a duration of 800us for 15 cycles. CV across the arrays was measured using the Cardio 2D software at each stimulus. Conduction velocities were generated from the software based on the time of stimulus application to measure electrical impulse across the array.

### CRISPR activation

Tracr RNAs targeting the *SCN5A* promoter and *SCN10A* intronic enhancer were cloned into backbones with sgRNA scaffold (addgene#47108). iPSC-aCMs were co-transfected with individual plasmids with sgRNAs, dCas9-VP64-p65-Rta (addgene#63798) or dCas9 CBP/ p300(addgene # 61357) and a plasmid with a GFP expression cassette (addgene#40973). dCas9-VP64-p65-Rta was used for activation of SCN5A promoter while dCas9 CBP/ p300 was used for SCN10A enhancer activation. All transfections were done using Lipofectamine 3000 according to the manufacturer’s protocol. 48 hrs post-transfection, GFP-positive iPSC-aCMs were selected for sodium current measurements.

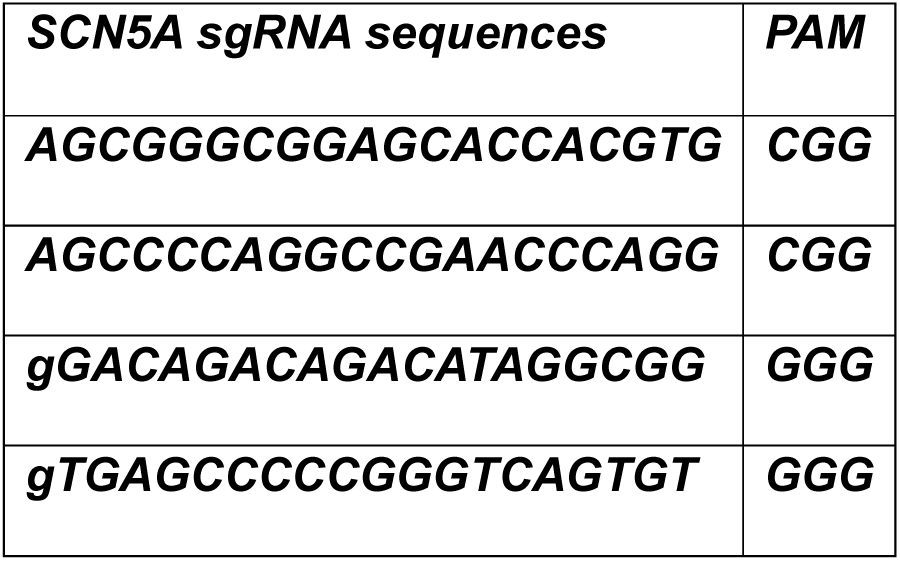

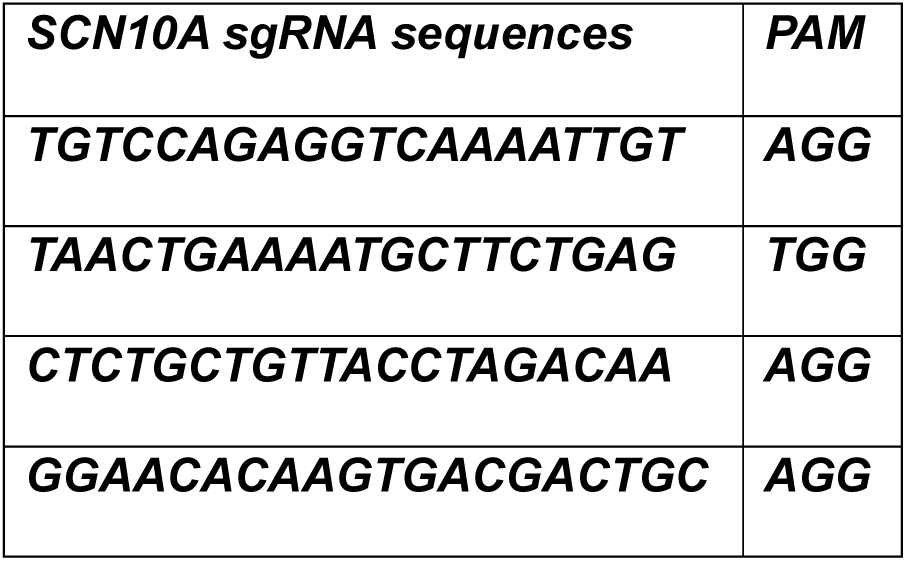

### RNA sequencing, ATAC sequencing, and Bioinformatic Analysis

RNA extraction and library preparation on triplicate samples of ∼ 1 million cells was done by the University of Chicago genome sequencing core. RNA was isolated using Trizol and QC-ed by bioAnalyzer (Agilent 2100); RNA-seq libraries were generated using an Illumina TruSEQ-like protocol (provided by Illumina). Libraries were QC-ed by BioAnalyzer and sequenced on the Illumina NovaSEQ-X using reagents and protocols provided by Illumina. Raw FASTQ files were uploaded to Biojupies (https://maayanlab.cloud/biojupies) for analysis. Gene enrichment analysis was performed using ShinyGO 0.81 and Ingenuity Pathway Analysis (IPA). IPA was also used for upstream regulator and transcription factor analysis.

For ATAC-seq, Triplicate samples of ∼ 500K single dissociated live iPSC-aCMs were provided to the Genomic Sequencing Core at Northwestern University for Library preparation. Briefly, the Cell membrane was permeabilized with nuclear lysis buffer (10 mM tris, 10 mM NaCl, 3 mM MgCl2, and 0.5% IGEPAL-630). Then, cells were resuspended in 50uL of transposase reaction mixture (22.5uL nuclease-free water, 25uL of TD buffer, and 2.5uL of TDE1 enzyme, Illumina #20034197). Transposition was carried out at 37°C for 22 min, followed by DNA purification with DNA Clean and Concentrator-5 (Zymo Research) according to the manufacturer’s recommendation. Following purification, library fragments were PCR amplified with Nextera XT v2 adapter primers. Multiplexed and pooled library was sequenced on the Novaseq X Plus with paired ends of 50 nucleotides per the manufacturer’s instructions. The library preparation and sequencing were done at the Northwestern University NUSeq facility core. The raw data was analyzed by the University of Illinois research informatics core.

### ATAC-seq Analysis Peak calling

Read alignments were first adjusted to account for TAC transposon binding: +4 bp for + strand alignments, −5 bp for - strand alignments. The open chromatin enrichment track was generated by first creating a bedGraph from the raw reads using bedtools genomcov^55^, then converted to bigWig using the UCSC tool bedGraphToBigWig ^56^. Tracks were normalized by the sum of alignment lengths of over 1 billion. The start position track was generated by taking just the first base of the alignment for + strand alignments or the last base of the alignment for - strand alignments, then creating bedGraph and bigWig tracks as for the open chromatin; tracks were normalized to the alignment count over 1 million. Open chromatin peaks were called using MACS2^57^ with -- nomodel set and no background provided; peaks with a score >5 were retained.

### Differential analysis of detected peaks

Differential analysis of quantitated peaks compared to genotype was performed using the software package edgeR on raw peak counts^58^ Before analysis, the data were filtered to remove any peaks that had less than 20 total counts summed across all samples. Data were normalized as counts per million, and an additional normalization factor was computed using the TMM algorithm. Statistical tests were performed using the “exactTest” function in edgeR. Adjusted p values (q values) were calculated using the Benjamini-Hochberg false discovery rate (FDR) correction^59^.Significantly enriched peaks were determined based on an FDR threshold of 5% (0.05).

#### RT-PCR

Total RNA was isolated from iPSC-aCMs using a Qiagen RNA extraction kit. Reverse transcription to synthesize cDNA was conducted using SuperScript III Reverse Transcriptase (Thermo Fisher Scientific). For the qPCR analysis, specific assays and primers were selected for target genes with glyceraldehyde 3-phosphate dehydrogenase (GAPDH) as the normalization reference gene. qPCR reactions were performed on an ABI QuantStudio 5 system (Applied Biosystems), using SYBR Green PCR Master Mix to accurately detect and quantify PCR amplification products.

Relative expression levels of the target genes were calculated using the ΔΔCt method by quantifying gene expression changes in the experimental samples relative to the control. For the ΔCt calculation, the cycling time (Ct) value of the target gene was subtracted from the Ct value of GAPDH in the same sample using ΔCt = Ct_target gene_ − Ct_reference gene_. The ΔΔCt value was then calculated using ΔΔCt = ΔCt_experimental_ *–* ΔCT_control_. The gene’s relative expression was calculated using relative gene expression = 2^−ΔΔCt^. **Primer pair for *SCN5A*-Forward-TCTCTATGGCAATCCACCCCA, Reverse-GAGGACATACAAGGCGTTGGT**

### Western Blotting

Protein lysates from iPSC-aCMs were isolated using 1× RIPA buffer. Based on previously published protocols. Each sample containing 50 μg of protein was subjected to SDS-PAGE gel electrophoresis. The resolved gels were then electro-transferred onto 0.2 μm PVDF membranes. After a 2-hour blocking step with 5% BSA, membranes were probed with specific antibodies for target proteins (Supplemental Table 3). Blots were developed using either anti-rabbit HRP or anti-mouse HRP and scanned with C280 imaging systems (Azure Biosystems). ImageJ software was used to determine protein expression levels. Antibody used for SCN5A-**68273-1-Ig SCN5A Mouse McAb from protein tech.**

### Immunofluorescence

iPSC-aCMs were washed with PBS, fixed with 4% paraformaldehyde at 37°C for 10 minutes, 0.1% Triton X-100 was used for permeabilization for 15 minutes, and blocked with 10% BSA for 1 hour. Primary antibody was diluted at 1:200 in 10% BSA and incubated in 10% BSA at 4°C overnight. The secondary antibody was incubated in 1:1000 dilution in 10% BSA for 1 hour. Primary antibodies utilized were rabbit polyclonal anti-cTnT (Abcam; ab45932) and mouse anti-sarcomeric anti–α-actinin (Abcam; EA-53 ab9465). Secondary antibodies used were goat anti-rabbit Alexa Fluor 488 (Abcam; ab150077) and goat anti-mouse Alexa Fluor 594 (Abcam; 150116).

Nuclei were stained using DAPI (Thermo Fisher Scientific). The cells were visualized using a Zeiss Laser Scanning confocal microscope (LSM 710) (META) objective and analyzed on ImageJ (NIH).

### Transmission Electron Microscopy

hiPSC-aCMs were fixed in Warmed (37°C) 2.5% glutaraldehyde in 0.1 M Sorenson’s Buffer for 60 min at room temperature, followed by gently scraping cells into a microcentrifuge tube prefilled with 2.5% glutaraldehyde in 0.1 M Sorenson’s buffer. Samples were centrifuged at 2500 g for 10 min at room temperature, and the pellet was dislodged and flipped with a hypodermic needle to ensure the fixative solution penetrated the entire sample. The pellet was kept at room temperature to fix for an additional 60 min. The 2.5% glutaraldehyde in 0.1 M Sorenson’s buffer solution was removed and replaced with 1% glutaraldehyde +4% paraformaldehyde in 0.1 M Sorenson’s buffer and stored at 4°C.

### Data Analysis and Statistics

All experiments were done in a minimum of three groups and repeated across at least three independent differentiations. Data was presented as mean ± SD. For data with normal distribution, nonparametric unpaired and 2-tailed Mann-Whitney U test was used to determine statistical significance between 2 groups and either 1-way or 2-way ANOVA for multiple groups with post hoc Bonferroni’s corrections. P < 0.05 was considered significant.

### UK Biobank and AoU PRS methods

The UK Biobank standard AF PRS (data field 26212) derived by Genomics PLC from external data sources was used^60,61^. The UK Biobank recruited over 500,000 participants aged 40 to 69 years old between 2006 and 2010 from multiple centers across the United Kingdom^62^. Ethics approval was obtained from National Research Ethics Service (reference 11/NW/0382). All participants provided written informed consent. Analyses were performed on the UK Biobank research analysis platform (application number 47602). The PRS for 420,196 participants was classified into 3 categories of genetic risk (low-risk [bottom quintile], high-risk [top quintile] and medium-risk [middle quintiles]. Logistic regression was used to assess the association of AF with PRS. Cox regression was used to calculate the hazard ratio (HR) for AF across PRS categories adjusting for the first 10 genetic PCs, sex, age and hypertension. Time to AF diagnosis (date at first diagnosis, data field 131350) was visualized with Kaplan Meier curves.

AoU Controlled Tier Dataset, version 7 was utilized in this study. AF cases were identified using OMOP Concept IDs: 313217, 321319, 3656119, 4064452, 4071896, 4108822, 4108832, 4117112, 4124693, 4141360, 4154290, 4163710, 4178584, 4199501, 4232495, 4232691, 4232697, 42872390, 44782442, 44783568, and 45768480. Whole-genome sequencing data from AoU were extracted and subjected to quality control using PLINK v1.9. This involved filtering out SNPs with a minor allele frequency (MAF) of < 0.01 and excluding SNPs with a genotyping rate < 95%.

SNP pruning was performed using PLINK with a 50 kb sliding window, shifted by 5 SNPs at each step, and an r² threshold of 0.2. The pruned SNPs were then used to compute genetic principal components (PCs) via PLINK. For Polygenic Risk Score (PRS) calculation, GWAS summary statistics for AF were obtained from a recent PRS analysis in individuals from the UK Biobank ^63^, and genomic coordinates were lifted over from hg19 to hg38 using the pyliftover tool (https://github.com/konstantint/pyliftover).

The PRS was classified into three categories of genetic risk: low-risk (bottom quintile), medium-risk (middle quintiles), and high-risk (top quintile). Survival analysis was conducted using Cox Proportional Hazards modeling, with age at diagnosis serving as the time-to-event variable.

## Data Availability

All data produced in the present study are available upon reasonable request to the authors.

## Videos of beating iPSC aCMs

**Supplementary Fig. 1:**
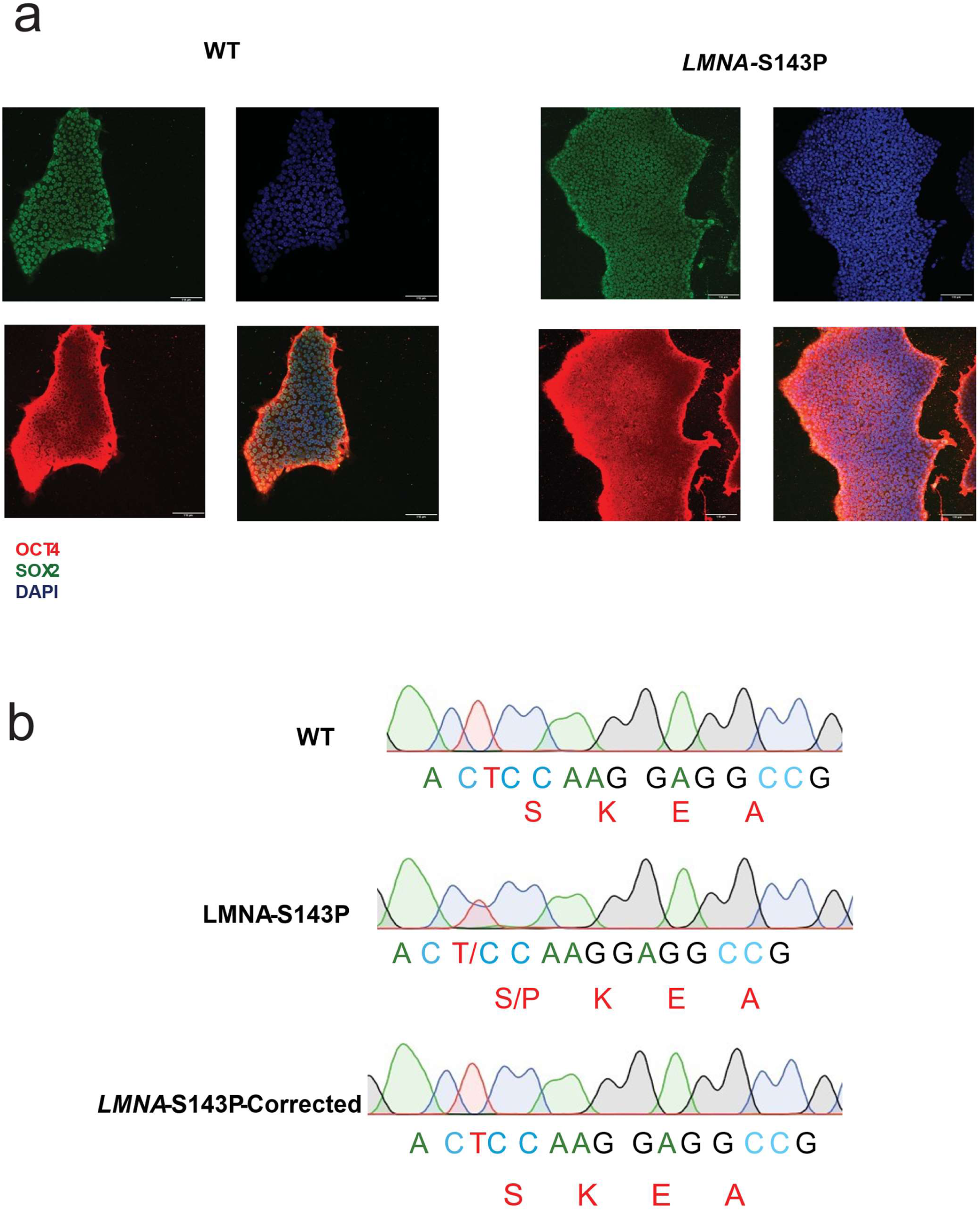
iPSC Validation and Sanger Sequencing. **a,** Immunostaining of pluripotency markers OCT4 and SOX2 in iPSCs. **b,** Sanger sequencing showing heterozygous missense variant at position 427 T>C resulting in an amino acid change from Serine to Proline.

**Supplementary Fig. 2:**
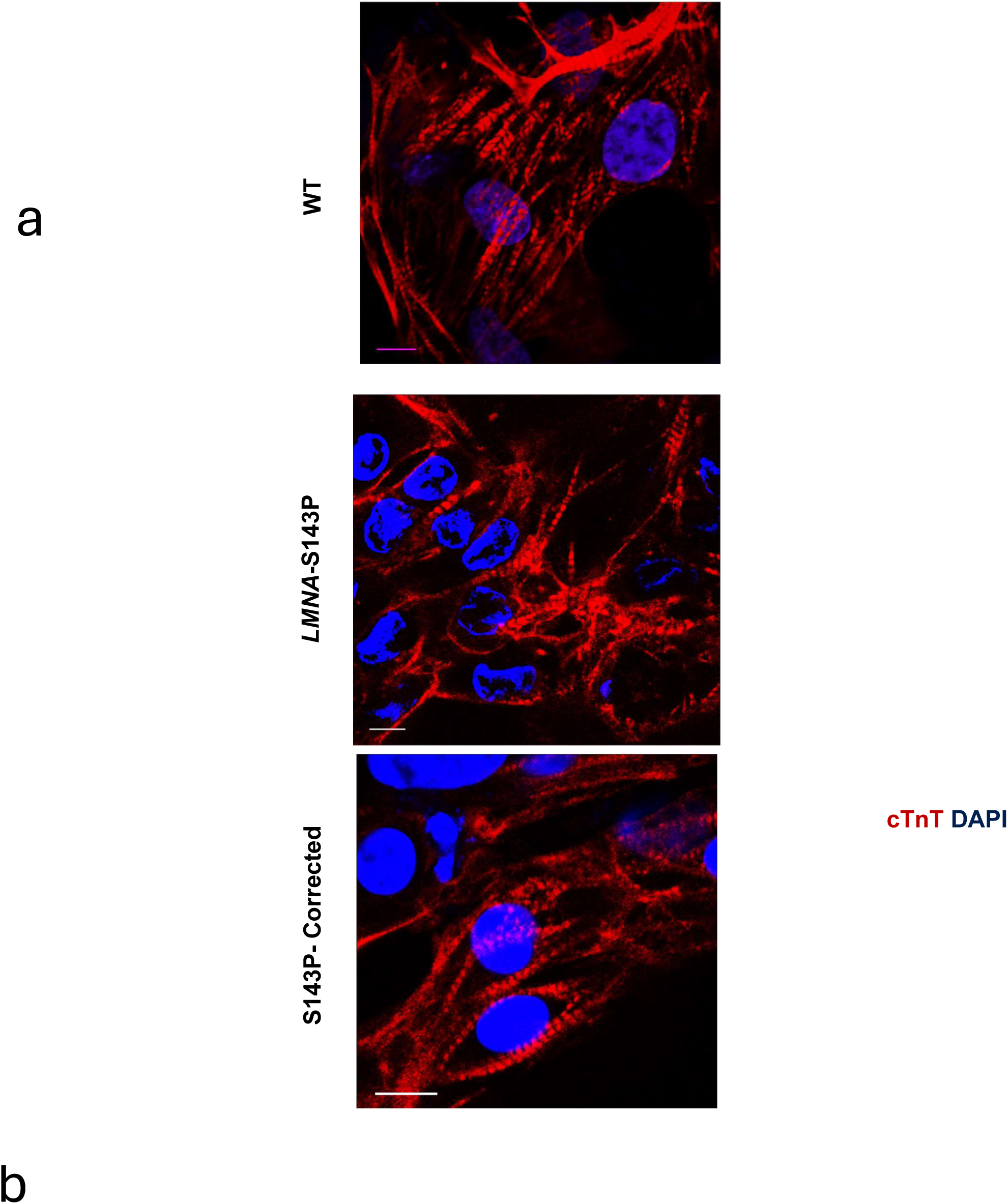
Immunostaining of iPSC-aCMs for cardiac troponin T. a, cTnT in red and DAPI in blue. **b,** Videos of beating iPSC-aCMs.

**Supplementary Fig. 3:**
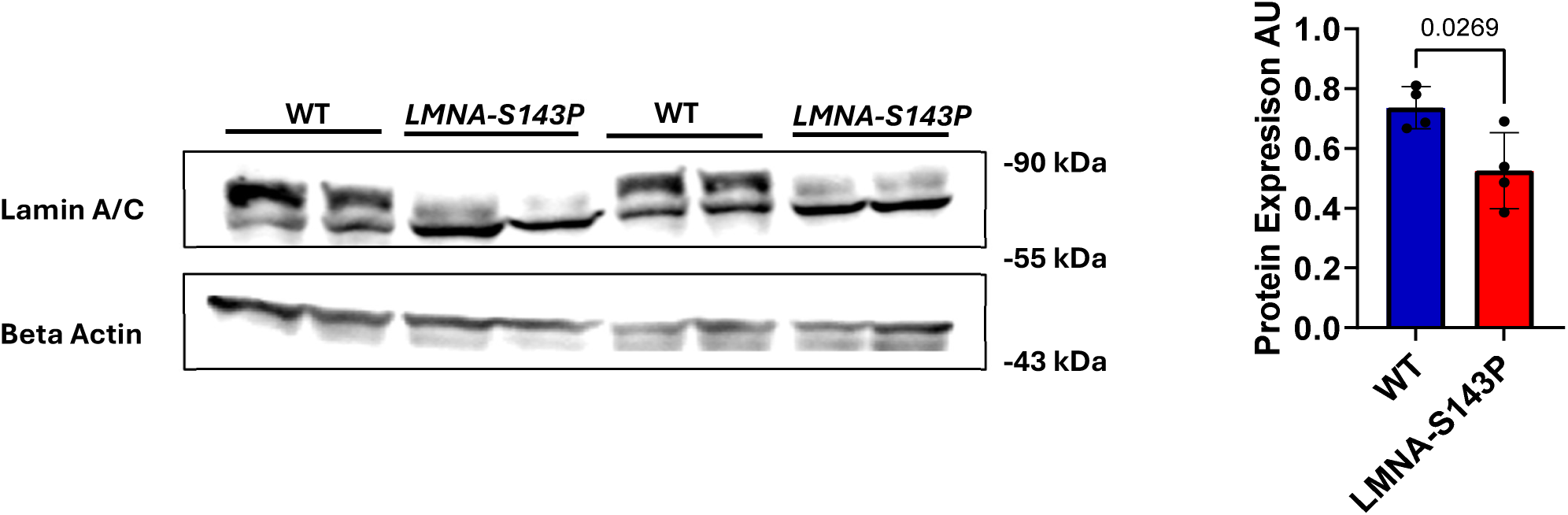
Protein expression of Lamin A/C isoforms in wild-type (WT) and *LMNA*-S143P iPSC-aCMs.

**Supplementary Fig. 4:**
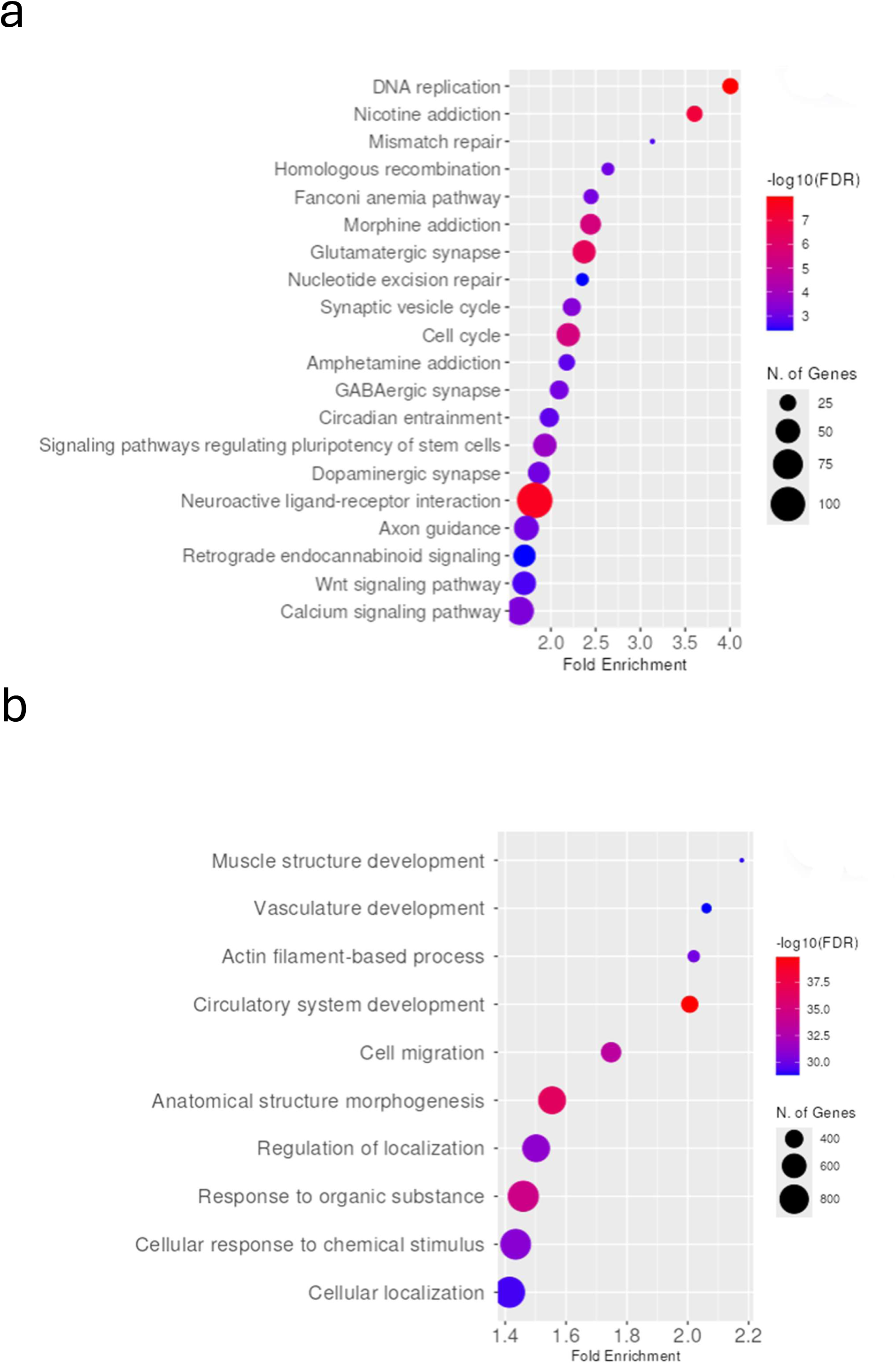
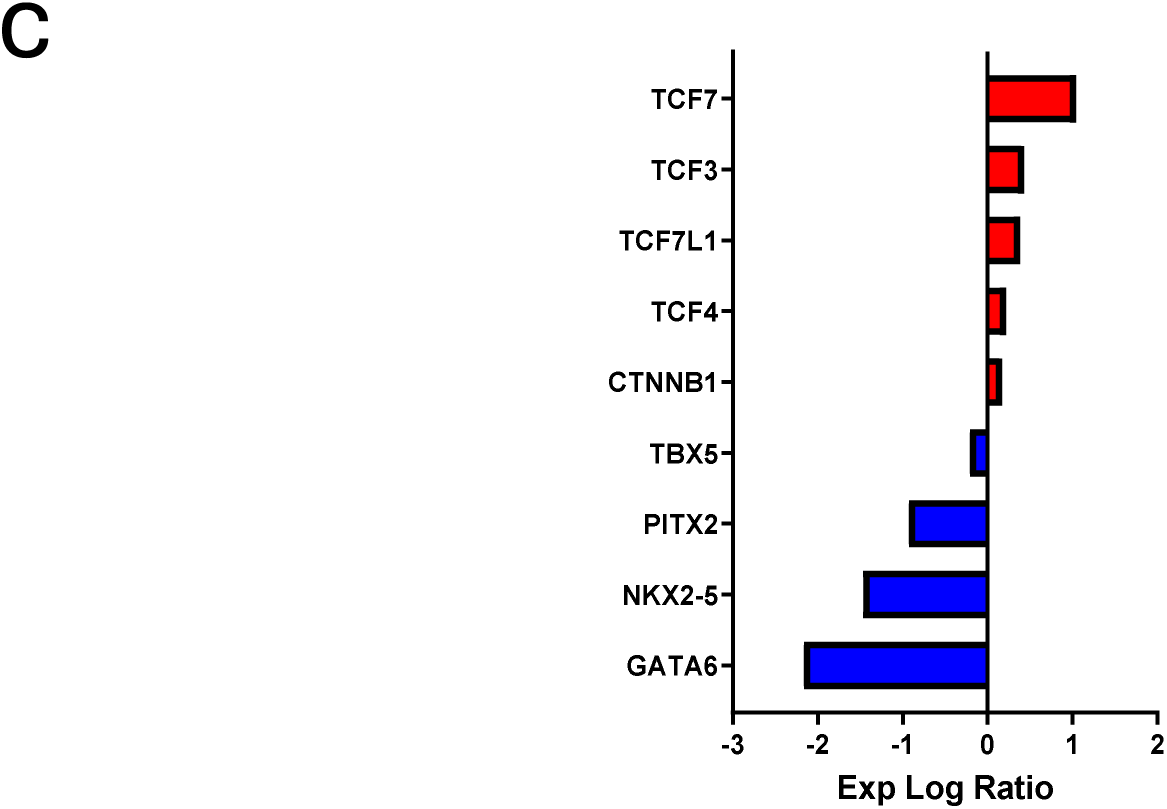
Transcriptomic changes in *LMNA*-S143P iPSC-aCMs. **a-b**, KEGG Pathway enrichment analysis of upregulated genes. **b,** GO enrichment analysis of downregulated genes. **c,** Transcription Factor enrichment analysis, e.g., Differential expression at Q < 0.05, log2FC > 0.

**Supplementary Fig. 5:**
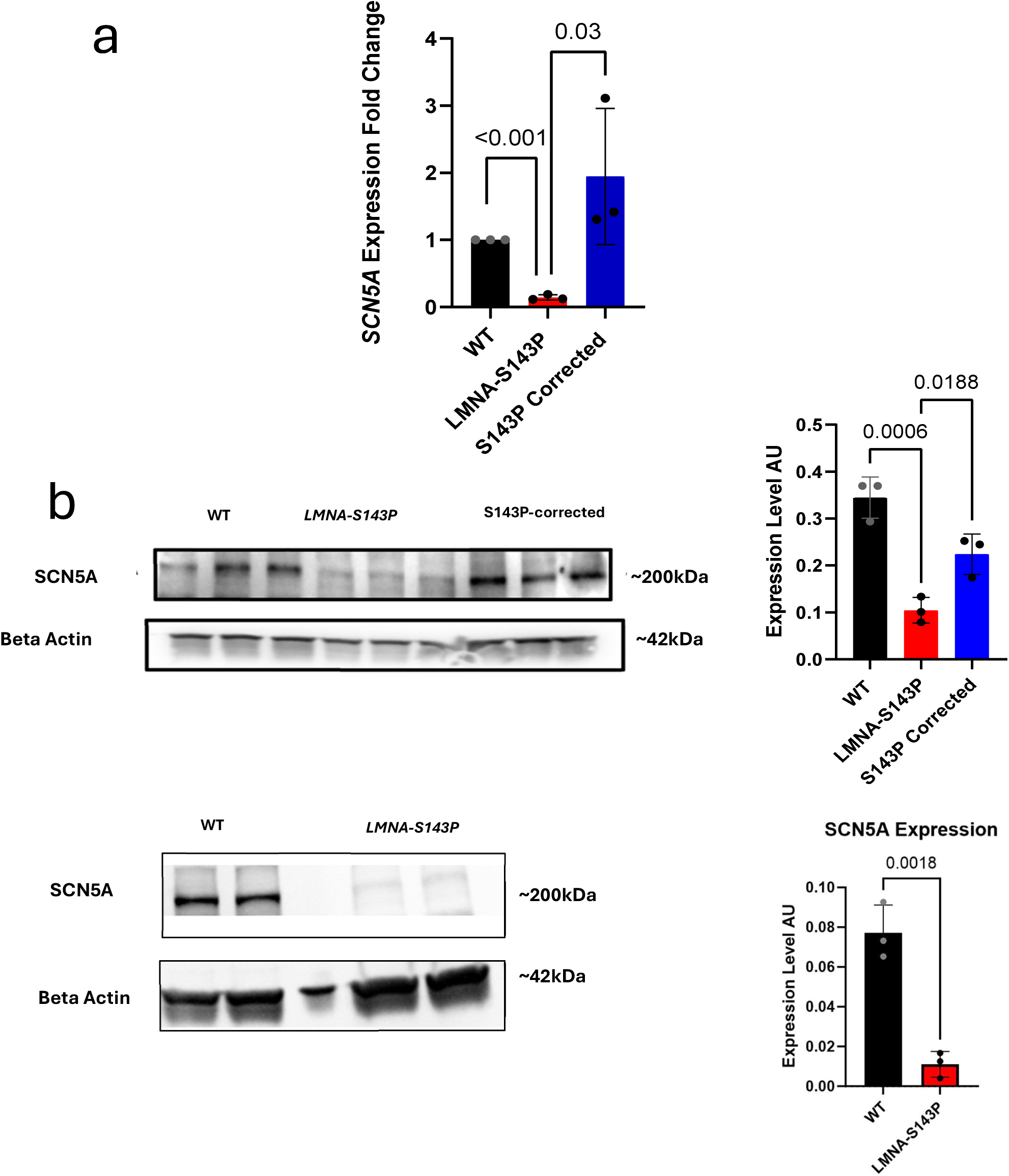
**a**. Reduced mRNA expression of *SCN5A* in *LMNA*-S143P iPSC-aCMs by qPCR. **b.** Reduced protein expression of *SCN5A* in *LMNA*-S143P iPSC-aCMs in samples from multiple differentiations.

**Supplementary Fig. 6:**
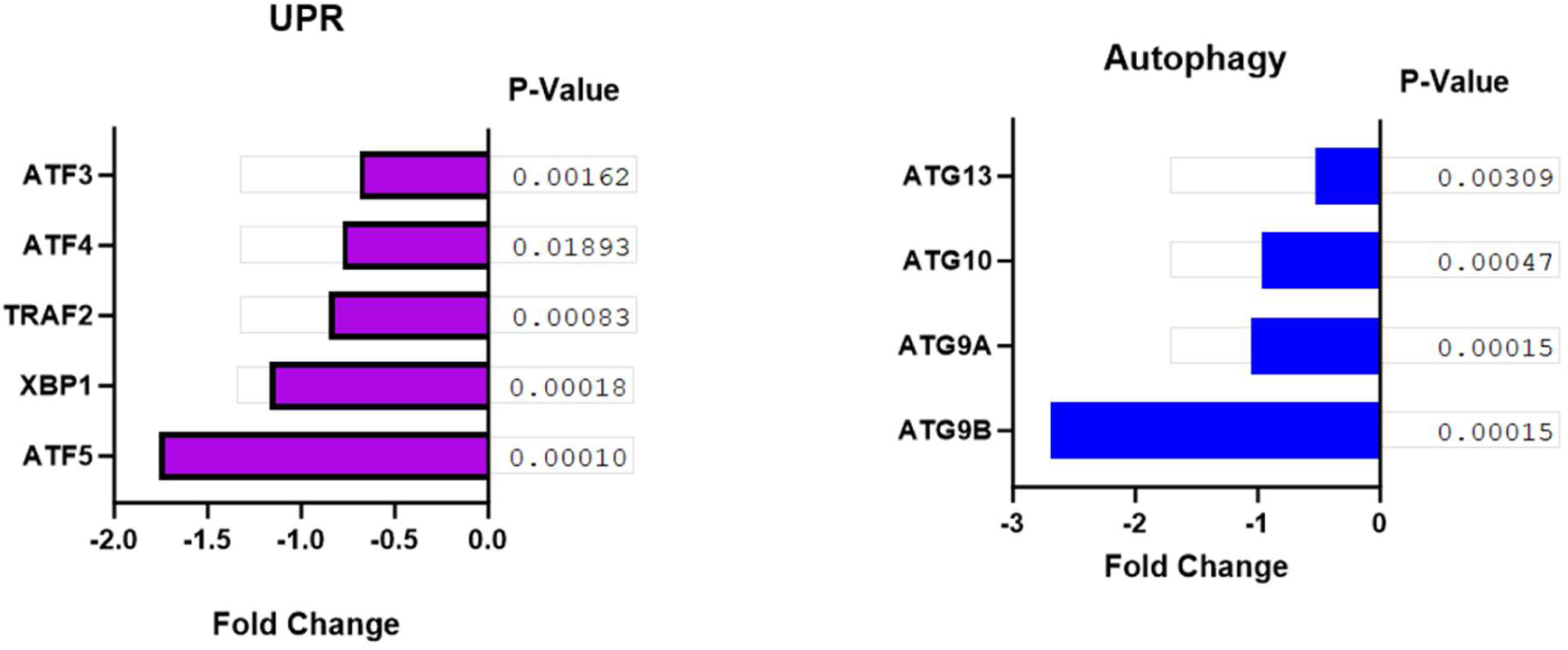
Expression fold change of genes involved in (a) Unfolded Protein Response (UPR) and (b) Autophagy (ATG) in *LMNA*-S143P iPSC-aCMs.

**Supplementary Fig. 7:**
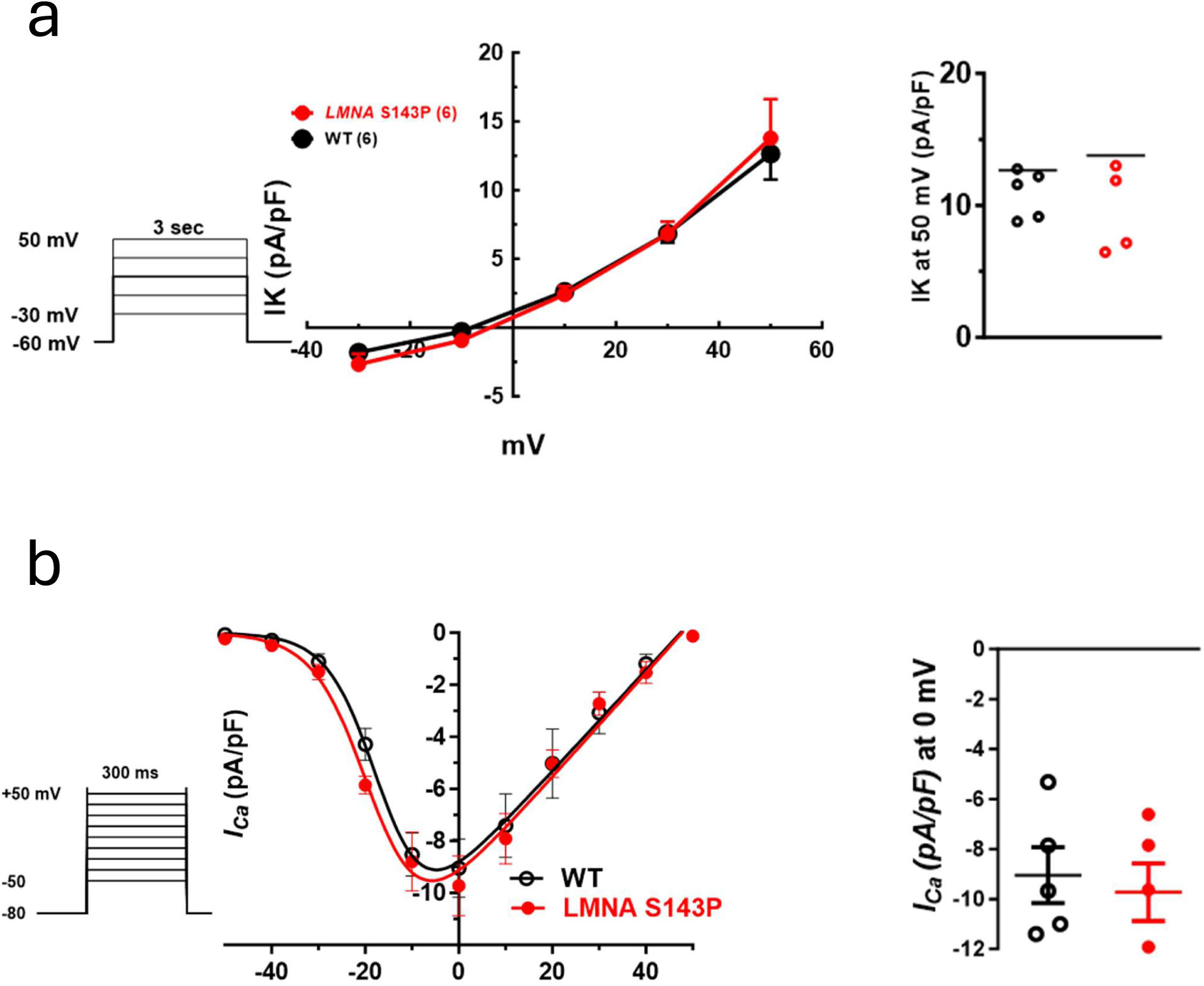
*LMNA*-S143P does not alter potassium (*I*_K_) or calcium (*I*_Ca,L_) currents. **a**, IV relationship for *I*_K_ in WT, *LMNA*-S143P iPSC-aCMs. **b,** IV relationship for *I*_CaL_ in the two groups.

**Supplementary Fig. 8:**
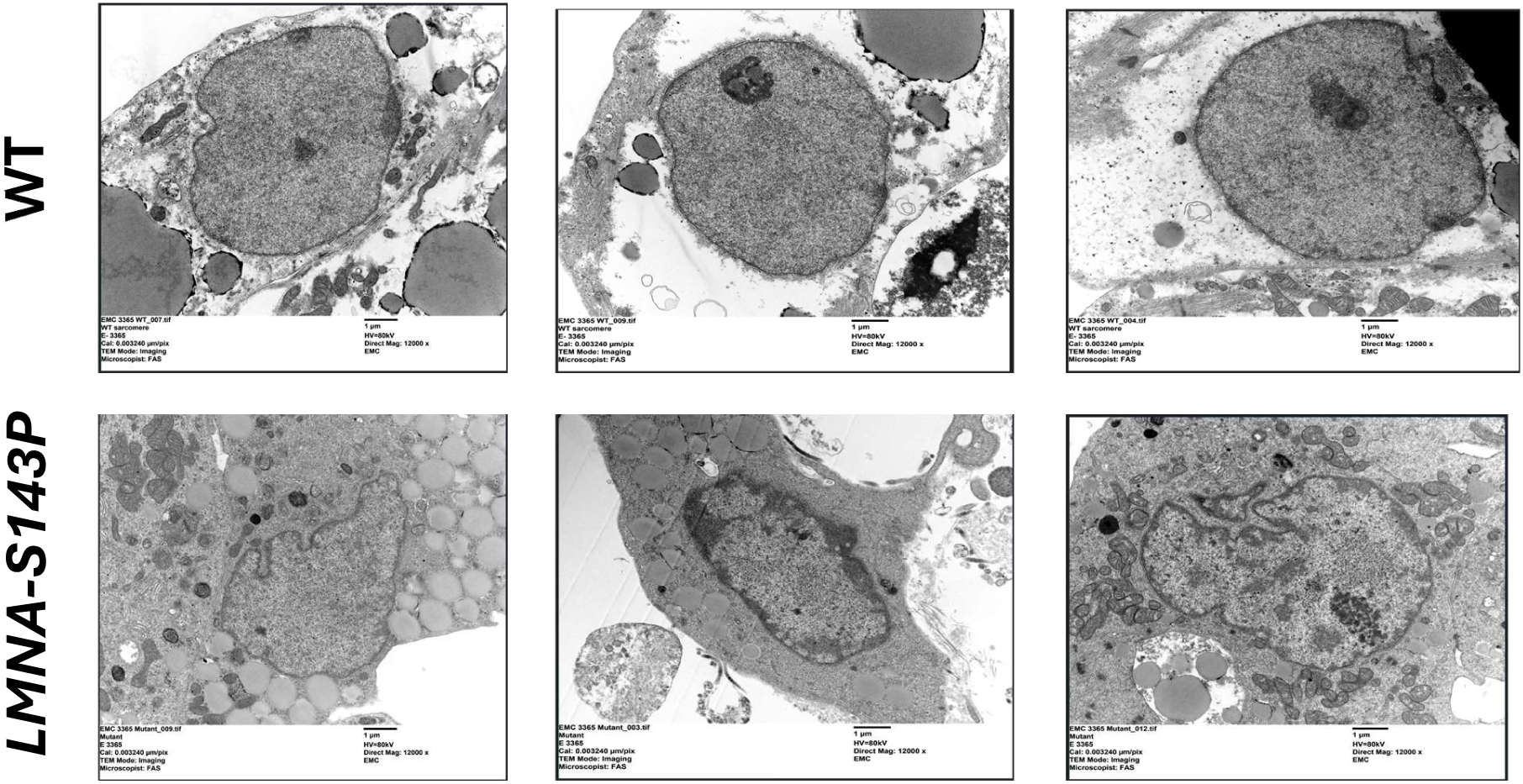
Transmission electron microscopy shows nuclear deformities in *LMNA*-S143P iPSC-aCMs.

**Supplementary Fig. 9.**
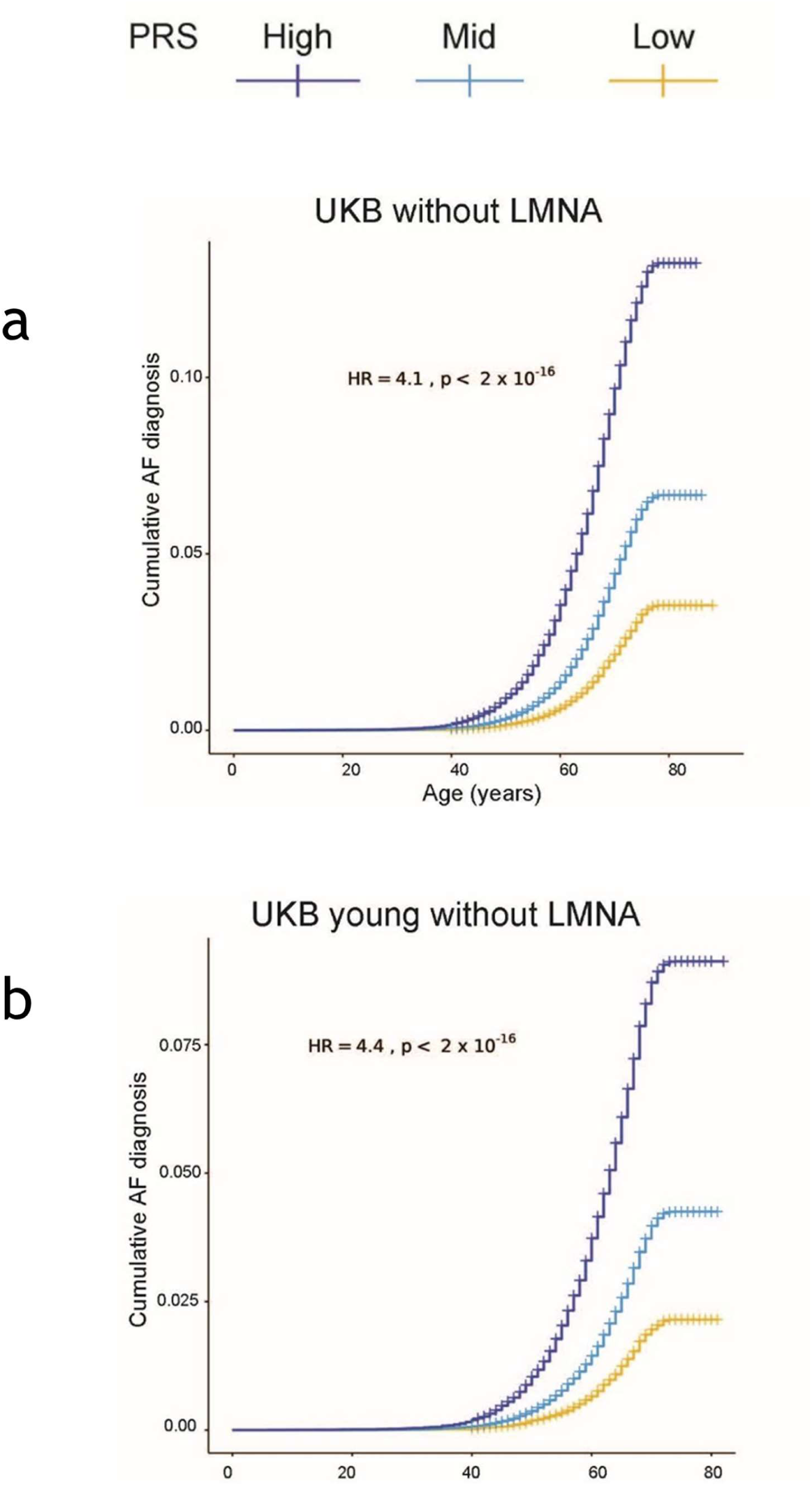
AF prevalence without PAV in *LMNA*. **a** Risk of AF in individuals who do not harbor PAVs in *LMNA* in the UK Biobank by PRS category (n = 419,087). A higher PRS is associated with a higher risk of AF in general. **b.** Risk of AF in young (<65 years) participants in the UKB who do not harbor PAVs in *LMNA* by PRS category.

**Supplementary Fig. 10:**
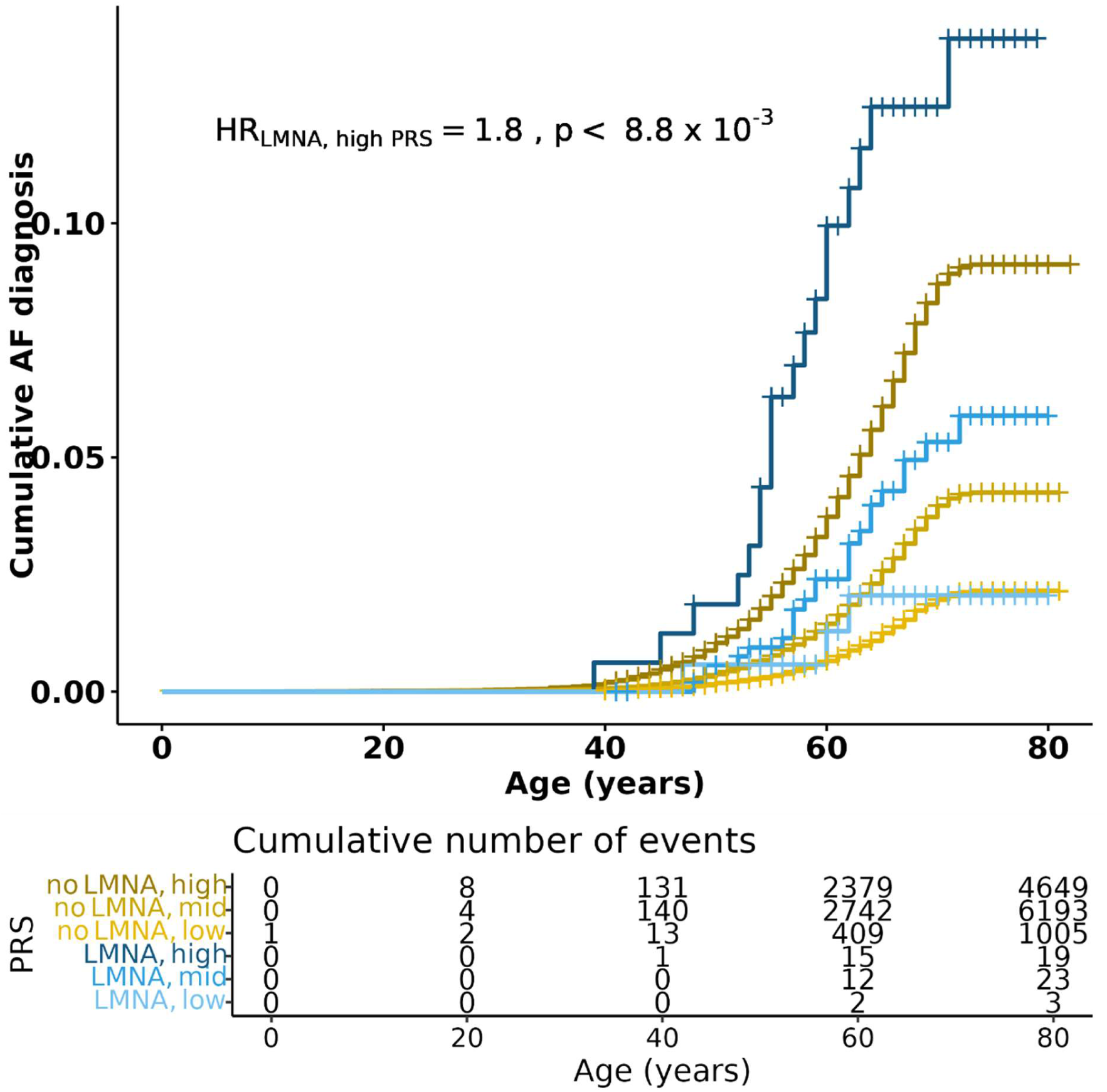
Risk of AF in the young (<65 years) UK Biobank participants using non*-LMNA* carriers with high PRS as the reference group. LMNA: LMNA carriers, nLMNA: non-LMNA carriers, low: low-risk AF PRS, mid: medium-risk AF PRS, high: high-risk AF PRS.

---

**Supplementary Table 1:** Overlap of AF risk loci and ATAC peaks in *LMNA*-S143P iPSC-aCMs.

| **rsID** | **rsID:Nearest_Gene(s)†** | **Chromosome** | **log2FC** | **p-value** |
| --- | --- | --- | --- | --- |
| **rs4896104** | ALDH8A1 | chr6 | -2.07327 | 0.004304 |
| **rs55734480** | DGKB | chr7 | -1.9301 | 0.008852 |
| **rs7172038** | NEO1 | chr15 | -1.73881 | 0.009063 |
| **rs6801957** | SCN10A | chr3 | -1.40184 | 0.00035 |
| **rs1769758** | ZMIZ1 | chr10 | -1.38549 | 3.73E-05 |
| **rs2540949** | CEP68 | chr2 | 0.634214 | 0.000908 |
| **rs12640611** | WDR1 | chr4 | 2.067337 | 8.50E-09 |
| **rs1886512** | KLF12 | chr13 | 2.179281 | 0.002199 |

**Supplementary Table 2:** Risk of AF in UK Biobank participants who do not harbor a PAV in *LMNA* by PRS category (n=419,087).

|  | No AF | Has AF | HR (p-value) |
| --- | --- | --- | --- |
| Low PRS (bottom 20%) | 81246 | 1881 | 1.00 |
| Mid PRS (20-80%) | 240175 | 11178 | 1.94 (p < 2 x 10<sup>-16</sup>) |
| High PRS (top 20%) | 76749 | 7860 | 4.11 (p < 2 x 10<sup>-16</sup>) |

**Supplementary Table 3:** Risk of AF in carriers of PAVs in *LMNA* in the UK Biobank by PRS category (n=1,109).

|  | No AF | Has AF | HR (p-value) |
| --- | --- | --- | --- |
| Low PRS (bottom 20%) | 216 | 5 | 1.00 |
| Mid PRS (20-80%) | 617 | 52 | 3.54 (p = 7.49 x 10<sup>-3</sup>) |
| High PRS (Top 20%) | 187 | 32 | 7.18 (p = 5.15 x 10<sup>-5</sup>) |

**Supplementary Table 4:** Risk of AF in young (<65 years) in *LMNA* PAV carriers compared to non-*LMNA* carriers.

|  | No AF | Has AF | HR (p-value) |
| --- | --- | --- | --- |
| Non-*LMNA* carriers | 320470 | 11847 | 1.56 (p = 3.01 x 10<sup>-3</sup>) |
| *LMNA* carriers | 823 | 45 |  |

**Supplementary Table 5:** Risk of AF in the young (< 65 yo) UK Biobank non-*LMNA* carriers by PRS category.

|  | No AF | Has AF | HR (p-value) |
| --- | --- | --- | --- |
| Low PRS (bottom 20%) | 64842 | 1005 | 1.00 |
| Mid PRS (20-80%) | 193150 | 6193 | 1.96 (p < 2 x 10<sup>-16</sup>) |
| High PRS (top 20%) | 62478 | 4649 | 4.4 (p < 2 x 10<sup>-16</sup>) |

**Supplementary Table 6:** Risk of AF in young (<65 years) in UK Biobank PAVs in LMNA carriers by PRS category.

|  | No AF | Has AF | HR (p-value) |
| --- | --- | --- | --- |
| Low PRS (bottom 20%) | 170 | 3 | 1.00 |
| Mid PRS (20-80%) | 511 | 23 | 3.08 (p = 7.11 x 10<sup>-2</sup>) |
| High PRS (top 20%) | 142 | 19 | 9.31 (p = 4.73 x 10<sup>-4</sup>) |

**Supplementary Table 7:** Risk of AF in UK Biobank P/LP LMNA carriers by PRS category.

|  | No AF | Has AF |
| --- | --- | --- |
| Low PRS (bottom 20%) | 5 | 0 |
| Mid PRS (20-80%) | 17 | 2 |
| High PRS (top 20%) | 9 | 4 |

## Notes

### Competing Interest Statement

The authors have declared no competing interest.

### Funding Statement

This study was supported by National Institutes of Health (NIH) R01 HL148444 (DD), NIH T32GM145767 (DD), NIH T32 HL139439 (DD), VA I01 BX004268 (DD), and the American Heart Association (AHA) 23PRE1010268 (AO).

### Author Declarations

The University of Illinois Chicago Institutional Review Board approved this protocol

### Summary of Updates

Some of the labels in Figure 1 were omitted in the initial version. The revised version has the updated figure with all labels included.

